# Toward clinical implementation of a metabolic blood biomarker for Parkinson’s disease differential diagnosis

**DOI:** 10.64898/2026.04.02.26349497

**Authors:** Vanille Millasseau, David Mallet, Sebastien Carnicella, Emmanuel L. Barbier, Mathilde Sauvee, Audrey Le Gouellec, Claire Cannet, Nils Pompe, Sabrina Boulet, Florence Fauvelle

**Author notes:** These authors contributed equally to this work (co-first authors) (Vanille Millasseau, David Mallet). These authors contributed equally to this work (co–senior authors) (Sabrina Boulet, Florence Fauvelle).

## Abstract

Parkinson’s disease (PD) diagnosis remains delayed and suboptimally accurate, largely due to clinical overlap with atypical parkinsonian syndromes and the lack of reliable biomarkers. In this context, we recently patented a 6-metabolites blood biomarker (6M-BB) for PD diagnosis versus healthy controls (HC), using high field nuclear magnetic resonance (NMR). Here, we evaluated its performance for the differential diagnosis of PD and its translation to clinical In-Vitro Diagnosis (IVDr) NMR platform. Patient serum samples from *de novo* PD (n=30), multiple system atrophy (MSA, n=30), progressive supranuclear palsy (PSP, n=30), Alzheimer’s disease (AD, n=33), and healthy individuals (n=29), were profiled by proton NMR both at 950 MHz and on an IVDr 600 MHz system. First, we reproduced the 6M-BB performance for PD vs. HC with 87.9% accuracy, but above all, we obtained an overall accuracy of 82.6% across all groups including MSA/PSP. Secondly, the 6M-BB refit using IVDr data achieved an overall accuracy of 77%. Adding VLDL-5 free cholesterol (V5FC) and citrate markedly increased accuracy to 95% for PD vs. HC and 85% vs. MSA/PSP. To conclude, in addition to independently validate the 6M-BB with a new PD/HC cohort, we demonstrated here its ability to efficiently distinguish PD from other confounding parkinsonian syndromes at *de novo* stage, thereby addressing a major challenge in the early clinical diagnosis of the disease. Its successful transfer to a fully automated, standardized IVDr system, with gains from V5FC and citrate, supports the feasibility and promising potential for clinical implementation, justifying future prospective multicenter studies.

**One Sentence Summary:** We offer a clinically deployable blood biomarker that addresses the major challenge of distinguishing Parkinson’s disease from other parkinsonism.

## INTRODUCTION

Parkinson’s disease (PD) is a progressive and currently incurable neurodegenerative disorder that affects more than 10 million people worldwide with prevalence having risen by more than 2.5-fold in recent decades (*1, 2*). It is characterized by a progressive loss of dopaminergic neurons in the substantia nigra pars compacta and by intracellular inclusions enriched in aggregated mutated α-synuclein (αSyn)(*3, 4*). Consistent with this ongoing neurodegeneration, the clinical presentation evolves gradually over years, from a prodromal phase dominated by nonspecific, non-motor symptoms (*5, 6*), to an overt motor stage that typically appears after approximately 40% to 60% loss of dopaminergic neurons (*7*).

To date, PD diagnosis remains clinical and relies primarily on the identification of the characteristic motor symptoms, with the *de novo* stage representing the earliest clinical phase in which newly diagnosed patients have not yet received dopaminergic treatment. In addition, this diagnosis often requires revisions over the course of the disease based on symptom evolution and response to anti-parkinsonian therapy—such as L-DOPA (*8–10*)—, making diagnosis accuracy highly dependent on both disease stage and clinician’s expertise (*9, 11*). Relevantly, post-mortem neuropathological studies—the only accepted reference standard for PD confirmation—have revealed misdiagnosis in up to one-third of PD *de novo* cases (*10, 11*). As a result, despite advancements in criteria (*12*), PD diagnosis remains delayed and suboptimal, prolonging care pathways, increasing social and economic burden (*13*), and hindering the timely implementation of effective neuroprotective strategies.

This suboptimal diagnostic performance mainly stems from the overlap between PD manifestations and those of atypical parkinsonian syndromes—particularly Multiple System Atrophy (MSA) and Progressive Supranuclear Palsy (PSP) (*14–17*)—combined with the current lack of approved and reliable *in vivo* biomarkers (*8*). In this context, substantial efforts have been deployed to develop robust diagnostic tools and biomarkers based on objective biological criteria reflecting PD-related pathophysiological processes (*18, 19*). Among these, DaTSCAN (DAT-SPECT) remains the only validated technique to reinforce *in-vivo* PD diagnosis, although it cannot reliably differentiates PD from MSA or PSP (*8, 20*). Additionally, new MRI-based imaging modalities are promising but remain expensive and poorly suited for widespread screening (*21, 22*). In response to the need for simple, cost-effective, and non-invasive diagnostic tools, molecular bio-fluid biomarkers have been increasingly explored (*23*). In particular, the cerebrospinal-fluid αSyn seed amplification assay (αSyn-SAA) has shown excellent sensitivity for PD; however it remains invasive, constraining and not fully specific (*18, 24–26*), and serum αSyn measurements has so far failed to produce consistent results (*27*). In parallel, blood neurofilament light chain (NfL) assays have shown potential for PD differential diagnosis and disease monitoring (*28, 29*), although its sensitivity remains limited in early stages (*29–31*).

Taken together, available diagnostic approaches—whether classical or emerging—are hindered either by limited specificity or by the invasiveness and the logistical burden of the current sampling and analytical procedures. This has led to growing interest in omics-based strategies (*23, 25, 32*), and in particular metabolomics. Indeed, given that PD is a multifactorial pathology resulting from a complex interplay of environmental and genetic factors (*33, 34*), metabolomics has emerged as a powerful approach to capture their combined influence, contributing to a better understanding of PD pathophysiology (*35, 36*) and the identification of candidate biomarkers (*37–40*).

In this regard, we recently identified a novel serum-based biomarker, composed of a specific combination of six metabolites—acetoacetate, betaine, β-hydroxybutyrate (BHB), creatine, pyruvate, and valine (patent PCT/FR2021/052035), following a large-scale from animal-to-human metabolomics study (*41*). Despite substantial heterogeneity in geography of *de novo* PD patients studied (two independent biobanks from USA and Italy), species used (rats, non-human primates and humans), and in PD induction models (toxin and human mutated αSyn models), this 6-metabolites blood biomarker (6M-BB), robustly distinguished *de novo* PD patients from healthy controls (HC) with 83% accuracy, a performance confirmed in an independent cohort. Furthermore, it successfully predicted PD in a prodromal-like animal model (*41*).

To further characterize the 6M-BB as PD-specific and a robust indicator of PD pathophysiology and to evaluate its clinical utility, we pursued two objectives: first, demonstrate its ability to distinguish PD from nonspecific neurodegeneration such as Alzheimer’s disease (AD) —as the leading neurodegenerative disorder worldwide—, and from confounding parkinsonian syndrome such as PSP and MSA; secondly, automate and standardize the analytical method to allow clinical applications. We showed a strong discriminatory performance, with an overall accuracy of 82.6% across all cases. Moreover, the implementation of the 6M-BB on a clinically deployable In-Vitro Diagnosis (IVDr) nuclear magnetic resonance (NMR) platform led to comparable diagnostic performance. Finally, we found that adding new metabolites and lipoprotein to the 6M-BB substantially improved its accuracy to 85%, further supporting its promise for future clinical use as a robust and scalable tool for PD differential diagnosis.

## RESULTS

### Cohort characteristics and sample quality control

The PD (n=30), HC (n=29), MSA (n=30), and PSP (n=30) groups showed similar age and sex ratio distributions. In the *de novo* PD group, the median time of the disease duration since diagnosis was 8 months (IQR: 6–22; range: 0-36 months), and their mean UPDRS-III score was 15.9 ± 10.6 (Table 1 and in Supplementary data, Table S1).

**Table 1.**
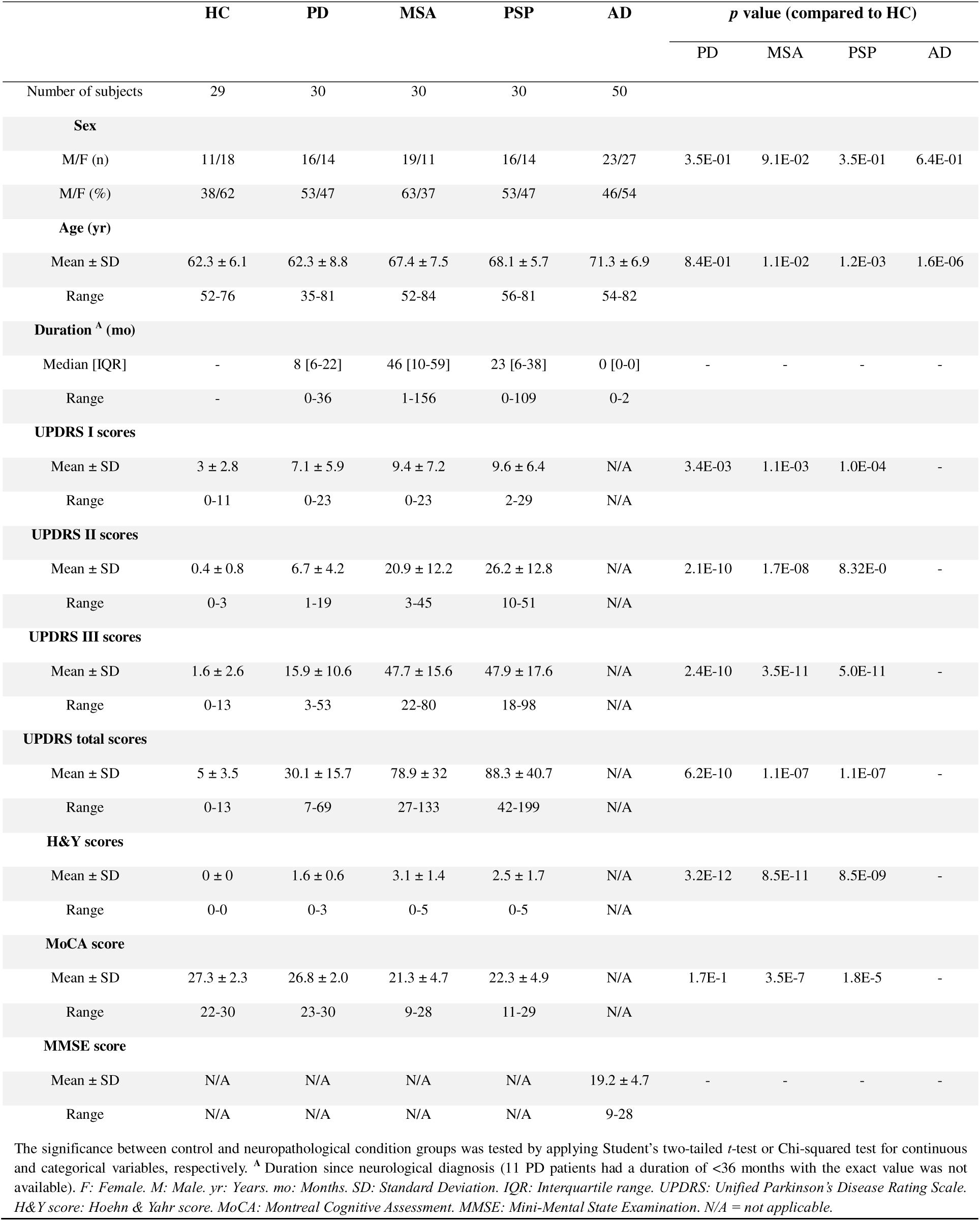
Demographic and clinical characteristics of the subjects included in this study.

Overall, the serum ^1^H NMR spectra were of high quality in terms of resolution and signal to noise ratio across all groups, enabling reliable quantification. In the 950 MHz ^1^H-NMR dataset, two spectra from the PDBP cohort (one HC and one MSA) were excluded due to excessive residual water signal. One individual (PSP) was excluded as a statistically significant outlier in acetoacetate and BHB amplitudes (Grubbs’ tests, p < 10 ^16^), which may reflect an atypical response to fasting.

### Confirmation of the 6-metabolites blood biomarker (6M-BB) performance with a new *de novo* PD cohort

First, in the score plot of the Orthogonal Partial Least Square Discriminant Analysis (OPLS-DA), we can observe that PD and HC groups are clearly separated, as expected (Figure 1A). This separation is supported by the model’s reasonable fit to the data matrix (R2Y=0.981) and a significant CV-ANOVA p-value (p=6.34×10^-3^). Strikingly, the six core metabolites composing the 6M-BB except the pyruvate were part of the most discriminating metabolites (Figure 1B). Consistently with our previous findings (*41*), betaine and valine levels were significantly decreased in PD patients compared to HC, whereas acetoacetate, BHB, and creatine levels were increased, although these changes were statistically significant for BHB only (Figure 1C). In parallel, four other metabolites (alanine, histidine, leucine, and lysine) showed significant alterations in PD samples (Supplementary data, Table S2), in accordance with our previous observations for leucine (*41*).

**Figure 1.**
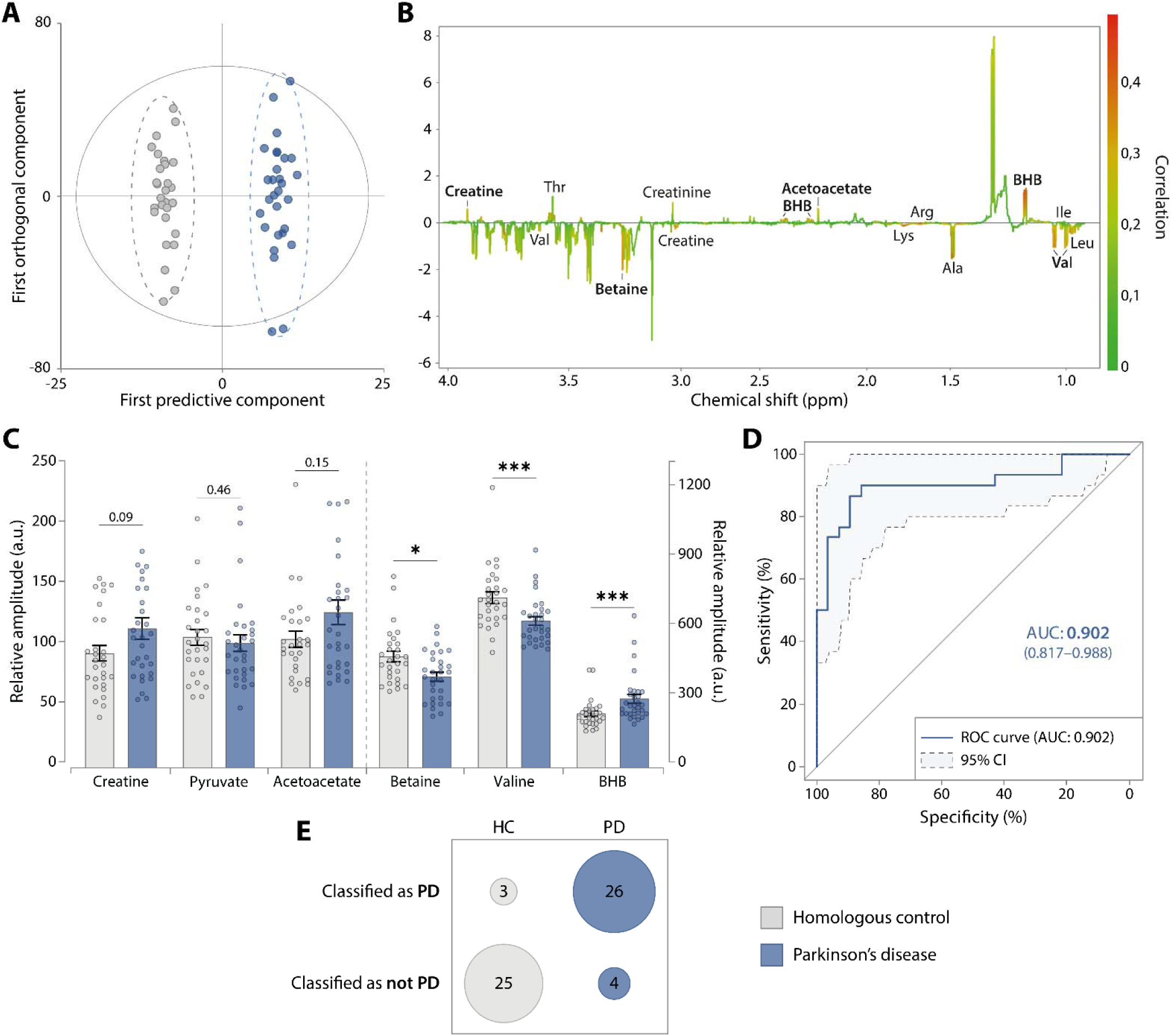
The 6M-BB accurately discriminates PD from HC. (**A** and **B**) Global multivariate analysis using OPLS-DA model built with PD (n=30, dark blue) and HC (n=28, grey) serum data. R2Y=0.981; Q2=0.433; 1 predictive and 5 orthogonal components; CV-ANOVA, *p*=6.34×10^-3^. (**A**) Scores plot of individuals based on first predictive/orthogonal components. A clear separation of the two groups is observed (**B**) Corresponding 1D loading plot (S-line). Each variable (bucket) is colored by its importance in the discrimination between the two groups, hot colors indicating highly correlated metabolites. Five of the six core metabolites (in bold) are among them. Positive/negative signs indicate their upregulation/downregulation in PD compared to HC (chemical shifts are listed in Supplementary data, Table S5). (**C**) Relative amplitude of the six core metabolites, showing that betaine, valine, and BHB significantly differ between PD and HC (Mean ± SEM. *p≤0.05; ***p≤0.001, Mann-Whitney tests). (**D**) ROC curve based on the 6M-BB logistic regression (*41*) with its 95% confident interval (light blue area within the dotted lines). AUC 0.902±0.09 indicates of a very robust classifier. (**E**) Classification table of PD and HC cases based on their 6M-BB scores (86.7% sensitivity, 89.3% specificity, 87.9% accuracy).

Secondly, to further evaluate the 6M-BB performance on this new independent cohort, we performed Receiver Operator Characteristic (ROC) curve analysis and constructed a classification contingency table using the previously described logistic regression model: *logit(P) = log(P/(1 − P)) = 42.79 + 0.17 BHB – 3.31 pyruvate – 12.01 valine + 7.06 acetoacetate + 3.14 creatine – 7.80 betaine* (*41*). The ROC curve yielded an Area Under the Curve (AUC) of 0.902 ± 0.09 (Figure 1D), consistent with our previous findings (*41*). These results correspond to an accuracy of 87.9% (sensitivity: 86.7%; specificity: 89.3%), demonstrating external high reproducibility of the 6M-BB for the classification of PD and HC samples (Figure 1E).

### Robust performance of the 6M-BB for PD differential diagnosis

Next, we evaluated whether the 6M-BB allows a differential diagnosis of PD against other neurodegenerative diseases, in particular atypical parkinsonian syndromes. We therefore analyzed samples from patients with AD and with PD-related disorders, MSA and PSP, and first compared global metabolic profiles across these groups alongside with PD and HC samples. The OPLS-DA model built with all cohorts data for purely descriptive purposes (Figure 2A), revealed in the score plot three main clusters: (i) a clearly separated cluster on the right side comprising AD patients, indicating that the AD metabolome is markedly distinct from the others; (ii) two partially overlapping clusters, with PD samples predominantly at the bottom and HC/ MSA/ PSP samples at the top, suggesting possibly a PD-specific metabolic profile. Consistently, the loading plot (Figure 2B) showed that acetoacetate, BHB, pyruvate, and valine ranked among the most discriminating metabolites between the PD and HC/MSA/PSP groups.

**Figure 2.**
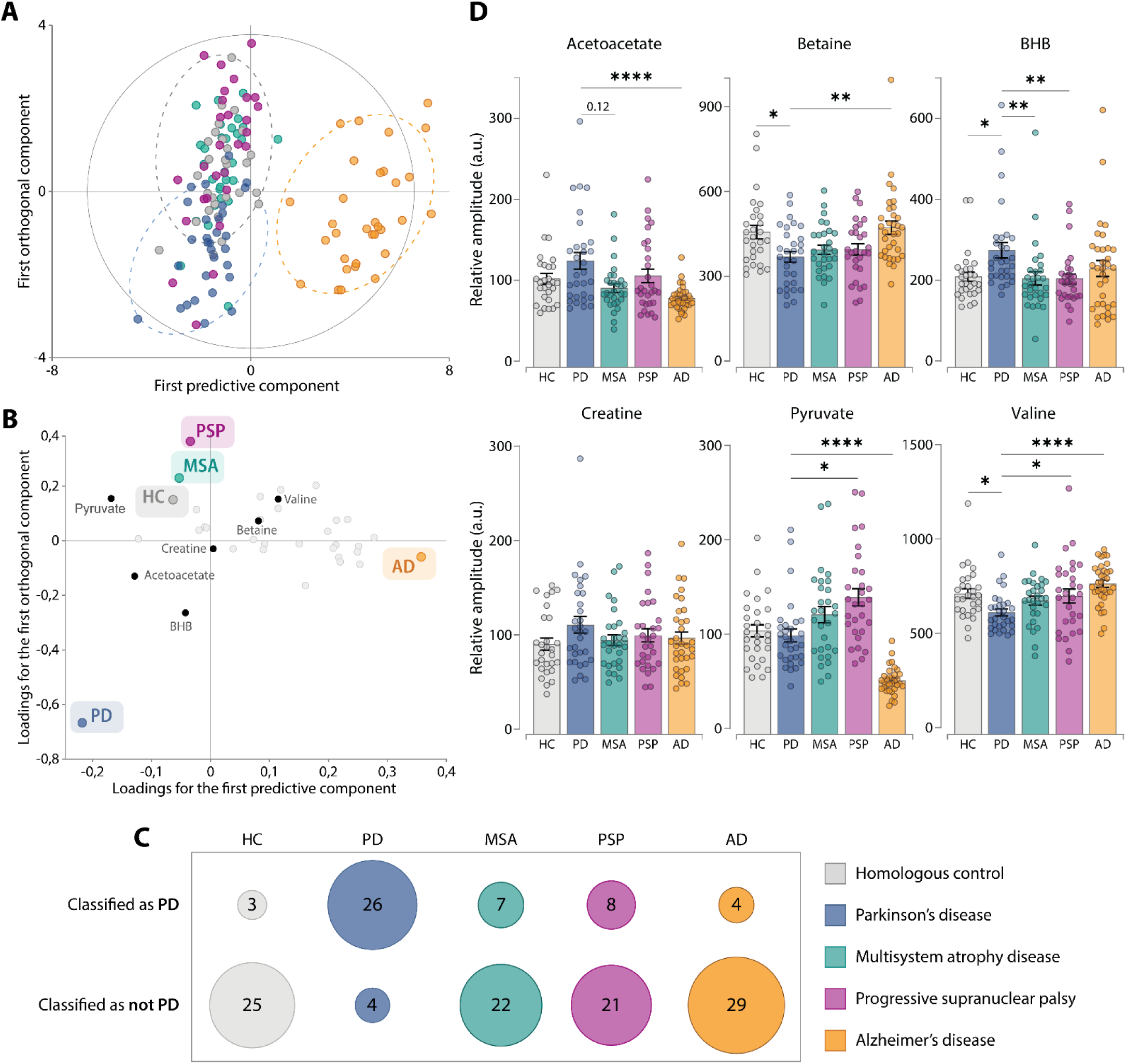
The 6M-BB accurately discriminates PD from other neurodegenerative diseases, including atypical parkinsonian syndromes. (**A** and **B**) Global multivariate analysis using OPLS-DA model built with relative metabolite amplitudes of all groups: HC (n=28, grey), PD (n=30, dark blue), MSA (n=29, turquoise), PSP (n=29, purple) and AD (n=33, orange). R2Y=0.296; Q2=0.248; 2 predictive and 1 orthogonal components; CV-ANOVA, *p*=6.29×10^-29^. (**A**) Score plot of individuals based on the first predictive/orthogonal components. Individuals are predominantly clustered in three groups: PD, HC/MSA/PSP and AD. (**B**) Corresponding 2D loading plot with the six core metabolites highlighted in bold. (**C**) Classification table of all cases based on their 6M-BB scores showing high PD differential diagnosis performance. Accuracies: MSA, 75.9%; PSP, 72.4%; and AD, 87.9%. Overall accuracy: 82.6% (81% considering only parkinsonian syndromes and HC). (**D**) Relative amplitudes of the six core metabolites in all cases, well aligned with 6M-BB classification results and PD discrimination (Mean ± SEM. *p ≤ 0.05; **p ≤ 0.01; ****p ≤ 0.0001, Kruskal-Wallis tests followed by Dunn’s post-hoc test for multiple comparisons against the PD group).

Accordingly, the 6M-BB was able to accurately classify over 87.9% of AD, 75.9% of MSA and 72.4% of PSP as not suffering from PD (Figure 2C). Taken together, the overall accuracy was 82.6%.

These classifications align with group-wise differences in the core metabolites (Figure 2D). Compared with PD, AD showed significantly lower levels of acetoacetate and pyruvate and higher levels of betaine and valine. Regarding PSP, BHB level was significantly lower, whereas pyruvate and valine levels were significantly higher than in PD. MSA exhibited similar variations, with BHB only reaching significance. Additionally, acetoacetate level also tended to decrease in MSA versus PD (*p*=0.12).

Overall, these results indicate that the 6M-BB captures PD-specific metabolic features and differentiates PD from clinically related atypical parkinsonian syndromes as well as from AD.

### Successful transfer of the 6M-BB to an IVDr platform for clinical use: the IVDr-6M-BB

To take a further step toward clinical applicability of the 6M-BB for PD diagnosis, we analyzed new aliquots from the serum samples already analyzed at 950 MHz on the Bruker Avance 600 MHz IVDr system, which allows standardized and automated pipeline with high-throughput analysis, detection, annotation and absolute quantification of 41 metabolites and 112 lipoprotein features. However, betaine quantification was currently not included in the method and was then performed separately, as described in Methods. Importantly, because the IVDr workflow outputs absolute concentrations, the original 6M-BB logistic regression coefficients, trained on relative concentrations, were not directly applicable. We therefore repeated the full metabolomics workflow at 600 MHz, from data acquisition to model construction, to derive new coefficients.

We first measured the levels of the six core metabolites. In PD, BHB and creatine were significantly increased and betaine decreased, associated with a trend toward an increase in acetoacetate (*p*=0.06) and no change in pyruvate (Figure 3A and Supplementary data, Table S3). Valine levels displayed a non-significant trend toward a decrease in PD, consistent with previous observations (Figure 3A and Supplementary data, Table S3). Overall, these results matched those observed at 950 MHz, supporting the transferability of the 6M-BB to the IVDr platform. The likelihood ratio test (p=2.03 × 10) confirmed that the model fit the data significantly better than the null model. We then computed IVDr-6M-BB scores (fitted values) and evaluated performance by ROC curve analyses (Figure 3B and 3C, respectively). The waterfall score plot (Figure 3B) demonstrates clear separation between HC and PD samples. Moreover, the training ROC curve yielded an AUC of 0.878 ± 0.09 (Figure 3C), which indicates strong discriminative ability with reasonable degree of uncertainty. Additionally, leave-one-out cross-validation (LOO-CV) gave an AUC slightly lower (0.808), as expected from optimism in the training estimate, yet still above 0.8. Overall, this indicates that the model presents good predictive power.

**Figure 3.**
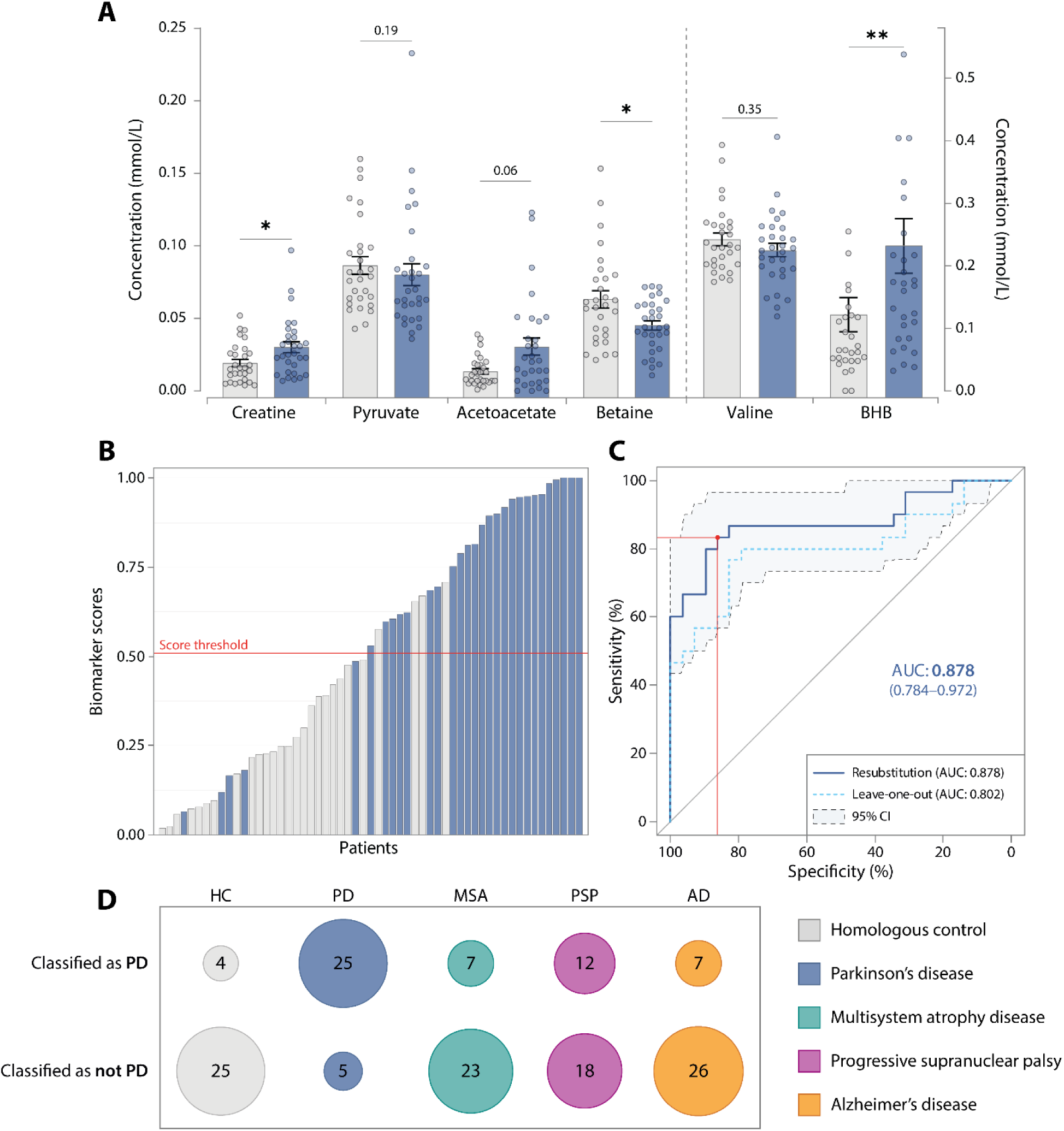
The IVDr-6M-BB accurately discriminates PD from HC and other neurodegenerative diseases, including atypical parkinsonian syndromes. (**A**) Absolute concentrations of the six core metabolites in HC (n=29, grey) and PD (n=30, dark blue), revealing similar variations to that observed previously. (Mean ± SEM. *p ≤ 0.05; **p ≤ 0.01, Mann-Whitney tests). (**B**) Waterfall plot of individuals IVDr-6M-BB scores. Cases above the threshold of 0.51 (red line) are classified as PD-positive, showing clear separation between true PD cases (dark blue) and controls (grey). (**C**) ROC curves based on the IVDr-6M-BB. Dark blue line: ROC curve obtained through resubstitution (AUC: 0.878 ± 0,09) with its 95% confident interval (light blue area within the dotted lines) and its optimal threshold (0.51; red line). Cyan line: ROC curve obtained through LOO-CV (AUC: 0.802). Both results indicate strong classifier robustness. (**D**) Classification table of HC, PD, MSA (n=30), PSP (n=30), and AD (n=33) cases based on their IVDr-6M-BB scores (threshold: 0.51), showing persistent good PD discrimination. Accuracies: HC, 86.2%; PD, 83.3%; MSA, 76.7%; PSP, 60%; and AD, 78.8%. Overall accuracy: 77% (76.5 % considering only parkinsonian syndromes and HC).

Importantly, the IVDr-6M-BB performance was comparable to that of the original 6M-BB, at least in terms of AUC (non-significant difference of 2.44%, DeLong’s test: *p*=0.7101). Moreover, using the optimal score threshold of 0.51, sensitivity and specificity reached 83.3% and 86.2% respectively (Figure 3B and 3C), similar to previously reported values.

Finally, with this classifier, 25/30 PD were true positives, and 25/29 HC, 23/30 MSA, 18/30 PSP, and 26/33 AD were true negatives (Figure 3D and Supplementary data, Figure S1). This corresponds to an overall accuracy of 77% (MSA accuracy: 76.7%; PSP accuracy: 60.0%; AD accuracy: 78.8%). Notably, in PSP group, the IVDr-6M-BB score correlated positively with disease duration since diagnosis (Spearman’s ρ=+0.44; p < 0.05) and PSP cases misclassified as “PD” had a significantly longer disease duration (median: 37.5 months) than correctly classified PSP cases (median: 20.5 months; Mann-Whitney, p=0.0224) (Supplementary data, Figure S2).

### Improvement of IVDr-6M-BB performance for PD diagnosis vs HC

The IVDr NMR pipeline provides extensive profiling of lipoprotein subclasses, in particular Very Low Density (VLDL), Low Density (LDL) and High Density (HDL) particles, offering a unique opportunity to identify new variables that further improve the IVDr-6M-BB for discriminating PD from HC samples. We therefore performed feature selection and model optimization using Least Absolute Shrinkage and Selection Operator (LASSO) regularization, combined with cross-validation. Seven metabolites or lipoproteins subclasses were retained: citrate; V2FC, V5FC, and H2FC (free cholesterol measures in VLDL/HDL subclasses); HDTG and H4TG (HDL-class triglycerides); and H3A2 (HDL-class apolipoprotein-A2). Adding each candidate one-by-one to the six core metabolites, we built logistic regression models and compared their metrics (Table 2A). Strikingly, four of the seven variables considerably enhanced the IVDr-6M-BB diagnostic performance: citrate and HDTG increased sensitivity by 7% (absolute), while V5FC and H2FC increased specificity by >10% (absolute). Among these, V5FC was the only variable significantly elevated specifically in PD patients, whereas citrate and HDTG showed slight upward trends in PD compared to HC, MSA, and PSP (Figure 4C and Supplementary data, Figure S3). Consistently, V5FC ranked among the most discriminating metabolites between PD and HC, appearing in the top-10 Variable Importance in the Projection (VIP) scores of the OPLS-DA model (Figure 4A and 4B).

**Figure 4.**
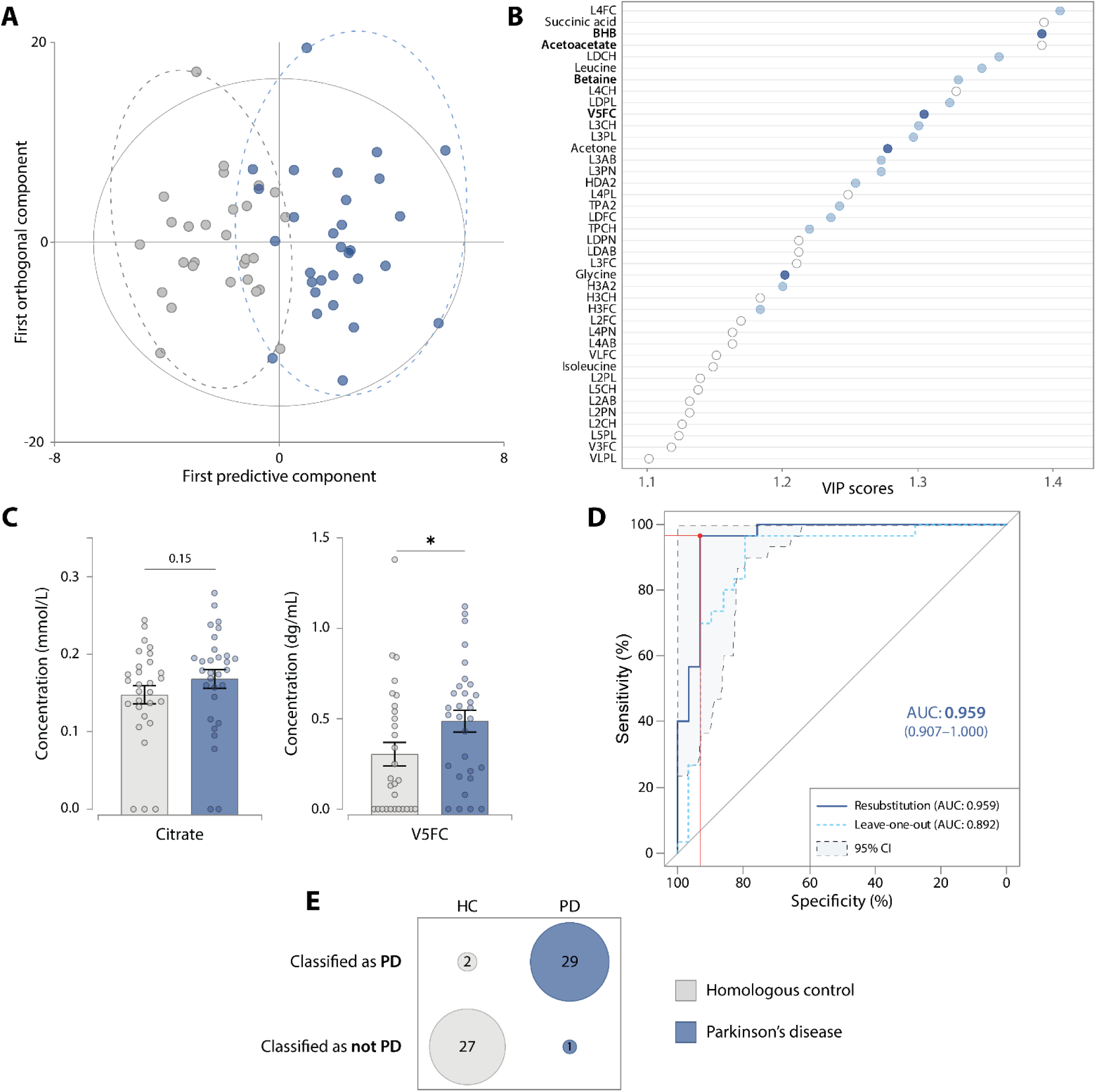
Citrate and V5FC improve the performance of the IVDr-6M-BB for PD versus HC classification. (**A** et **B**) Global multivariate analysis using OPLS-DA model built with HC (n=29, grey) and PD (n=30, dark blue) metabolites and lipoproteins data. R2Y=0.643; Q2=0.278; 1 predictive and 3 orthogonal components; CV-ANOVA, *p*=0.0314. (**A**) Scores plot of individuals based on first predictive/orthogonal components. A clear separation of the two groups is observed. (**B**) Top-40 VIP, revealing some of the core metabolites with V5FC (in bold) in the 10-top VIP. Dark blue/light blue: metabolites significantly increased/decreased in PD samples compared to HC (Mann-Whitney tests, detailed *p* values in Supplementary data, Table S2). (**C**) Citrate and V5FC concentrations, demonstrating dysregulation in PD (Mean ± SEM. *p ≤ 0.05, Mann-Whitney tests). (**D** and **E**) Performance of the logistic regression of the IVDr-6M-BB+citrate+V5FC, highlighting improved PD discrimination. (**D**) ROC curves analysis. Dark blue line: ROC curve obtained through resubstitution (AUC: 0,959 ± 0,05) with its 95% confident interval (light blue area within the dotted lines) and its optimal threshold (0.4937; red line). Cyan line: ROC curve obtained through LOO-CV (AUC: 0.892). (**E**) Classification table of cases based on their IVDr-6M-BB+citrate+V5FC scores (96.7% sensitivity, 93.1% specificity, 94.9% accuracy).

**Table 2.** Addition of new variables to improve the IVDr-6M-BB performance.

| Models – PD vs HC | AUC<br>(95%CI)<sup>c</sup> | Threshold | Sensitivity<br>(%) | Specificity<br>(%) | PPV<br>(%) | NPV<br>(%) | Accuracy<br>(%) | AUC-LOO<sup>d</sup> |
| --- | --- | --- | --- | --- | --- | --- | --- | --- |
| **Models with one added**<br>**candidate variables <sup>A</sup>** |  |  |  |  |  |  |  |  |
| IVDr-6M-BB + Citrate | 0.940 (0.88-1) | 0.471 | 90.00 | 86.21 | 87.10 | 89.29 | 88.14 | 0.870 |
| IVDr-6M-BB + H4TG | 0.932 (0.87-0.99) | 0.546 | 86.67 | 89.66 | 89.66 | 86.67 | 88.14 | 0.833 |
| IVDr-6M-BB + HDTG | 0.928 (0.86-1) | 0.428 | 90.00 | 89.66 | 90.00 | 89.66 | 89.83 | 0.841 |
| IVDr-6M-BB + H2FC | 0.926 (0.86-0.99) | 0.584 | 80.00 | 96.55 | 96.00 | 82.35 | 88.14 | 0.837 |
| IVDr-6M-BB + V5FC | 0.914 (0.84-0.99) | 0.646 | 80.00 | 96.55 | 96.00 | 82.35 | 88.14 | 0.818 |
| IVDr-6M-BB + H3A2 | 0.911 (0.84-0.99) | 0.499 | 86.67 | 86.21 | 86.67 | 86.21 | 86.44 | 0.832 |
| IVDr-6M-BB + V2FC | 0.906 (0.84-0.99) | 0.430 | 86.67 | 79.31 | 81.25 | 85.19 | 83.05 | 0.795 |
| IVDr-6M-BB | 0.878 (0.78-0.97) | 0.510 | 83.33 | 86.21 | 86.21 | 83.33 | 84.75 | 0.802 |
| **Models with multiple added**<br>**candidate variables <sup>B</sup>** |  |  |  |  |  |  |  |  |
| IVDr-6M-BB + V5FC + Citrate + HDTG | 0.99 (0.97-1) | 0.445 | 96.67 | 93.10 | 93.55 | 96.43 | 94.92 | 0.798 |
| IVDr-6M-BB + V5FC + Citrate | 0.959 (0.91-1) | 0.494 | 96.67 | 93.10 | 93.55 | 96.43 | 94.92 | 0.892 |
| IVDr-6M-BB + V5FC + HDTG | 0.938 (0.87-1) | 0.435 | 90.00 | 89.66 | 90.00 | 89.66 | 89.83 | 0.843 |

ROC curve analyses of the different logistic regression models based on PD (n=30) and HC (n=29), each including the 6 core metabolites (acetoacetate, BHB, betaine, creatine, pyruvate, valine) plus additional variables. Models are listed in descending order of AUC. <sup>A</sup>Addition of one of the seven LASSO-selected candidate variable. <sup>B</sup>Different combinations of the following variables: V5FC, HDTG and citrate. <sup>C</sup>AUC of ROC curve obtained through resubstitution with a 95% confidence interval. <sup>D</sup>AUC of ROC curve obtained through Leave-One-Out (LOO) cross-validation. *AUC: Area Under the Curve. CI: Confident Interval. PPV: positive predictive value; NPV: negative predictive value.*

Guided by these results, we performed models that combined the six core metabolites with V5FC (to enhance specificity), and either citrate, HDTG, or both (to improve sensitivity)(Table 2B). The best-performing models, which achieved nearly 95% overall accuracy, both included V5FC and citrate, and adding HDTG did not improve their performance. Taken together, augmenting the core metabolites panel with V5FC and citrate substantially improved IVDr-6M-BB performance. ROC curve analysis confirmed this enhancement, yielding and AUC of 0.959 ± 0.05 (Figure 4D). Although this model appears to be slightly overfitted, its LOO-CV AUC of almost 0.9 indicates strong discriminative ability. At the optimal threshold (0.4937), the model reached a ∼95% accuracy with 96.7% sensitivity and 93.1% specificity (Figure 4E).

### High performance of the improved IVDr-6M-BB for PD differential diagnosis

Subsequently, we evaluated whether adding V5FC and citrate to the IVDr-6M-BB improves differential diagnosis of PD from atypical parkinsonism syndromes, which is the main clinical issue that neurologists have to deal with. To this end, we built pairwise OPLS-DA models with either PD vs. MSA or PD vs. PSP, and plotted the top-30 VIP scores. The score plots showed clear clustering for both PD vs. MSA and PD vs. PSP (Figures 5A and 5C). Although the models had acceptable explanatory power (R²Y > 0.5), Q² <0.5 indicated limited predictive ability. Nevertheless, CV-ANOVA *p* values were below the significance threshold, indicating that the observed group separation is unlikely due to chance. Therefore, although this model should be interpreted with caution, the VIP score plots can provide insight into the variables driving the discrimination between PD and MSA/PSP samples. Notably, acetoacetate, BHB, and pyruvate remained important metabolites for distinguishing PD from MSA or PSP samples, being part of the top-10 VIP metabolites identified in both models (Figure 5B and 5D). Valine contributed also to the separation between PD and MSA (VIP score > 1). Strikingly, V5FC was the top-ranked VIP in both models, highlighting the central role of this lipoprotein subclass in distinguishing PD from both MSA and PSP.

**Figure 5.**
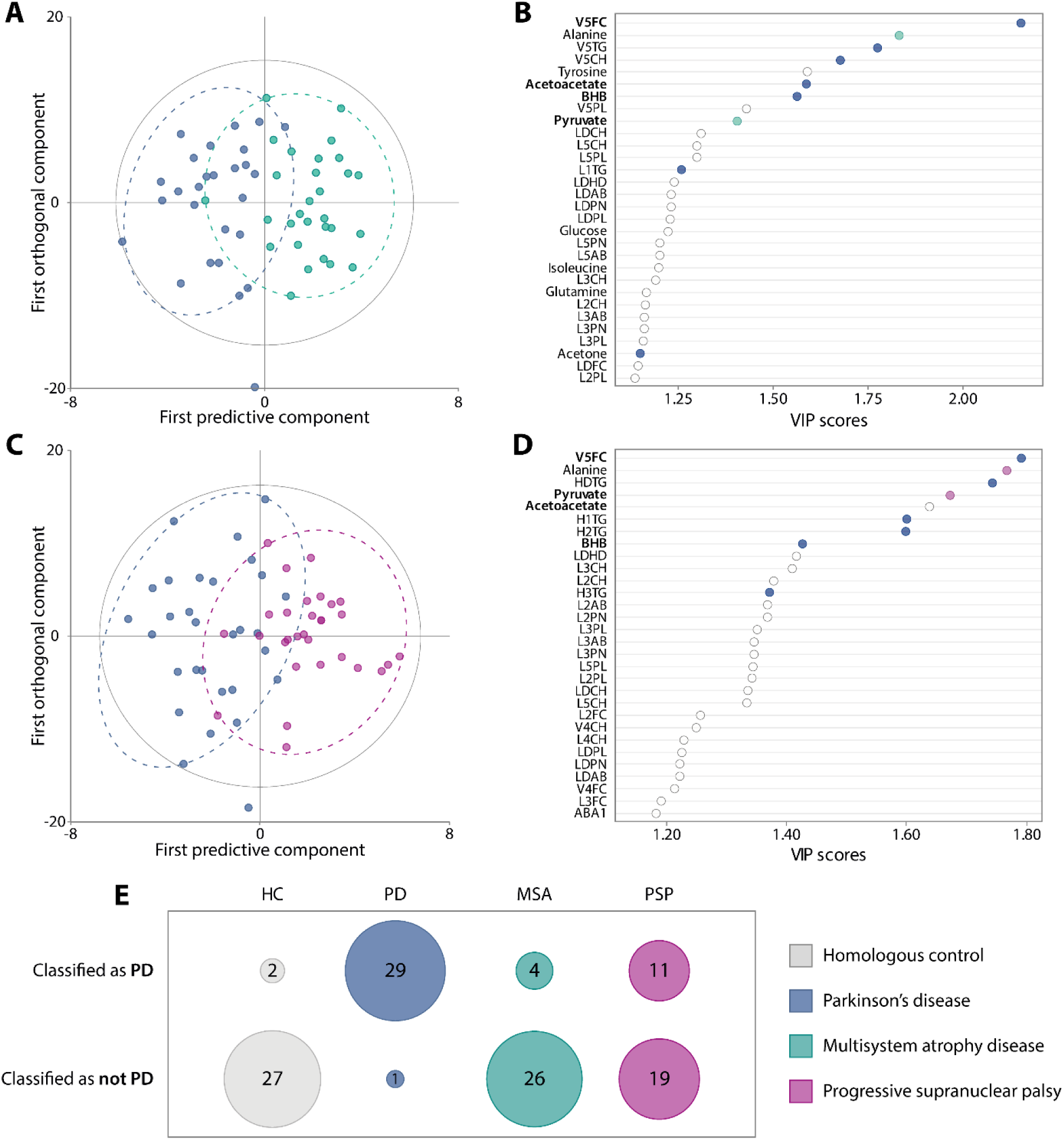
Citrate and V5FC improve the performance of the IVDr-6M-BB for differential diagnosis of PD against confounding parkinsonian syndromes. (**A-D**) Global multivariate analysis using OPLS-DA model built from (**A** and **B**) PD (n=30, dark blue) and MSA (n=30, turquoise), or (**C** and **D**) PD and PSP (n=30, purple) metabolite and lipoprotein data, showing clustering by case groups. PD-MSA model parameters: R2Y=0.644; Q2=0.311; 1 predictive and 3 orthogonal components; CV-ANOVA, *p*=0.01742. PD-PSP model parameter: R2Y=0.569; Q2=0.213; 1 predictive and 2 orthogonal components; CV-ANOVA, *p*= 0.04383. (**A**, **C**) Score plot of individuals based on the first predictive/orthogonal components. (**B**, **D**) Top-30 VIP plot, revealing some of the core metabolites with V5FC (in bold) in the 10-top VIP. Colors indicate direction of change: dark blue for metabolites increased in PD, turquoise/purple for metabolites increased in MSA/PSP (Mann-Whitney tests, detailed *p* values in Supplementary data, Table S2). (**E**) Classification table of HC (n=29), PD, MSA, and PSP cases based on their IVDr-6M-BB+citrate+V5FC scores (threshold: 0.4937), showing improved PD differential diagnosis performance. Accuracies: HC, 93.1%; PD, 96.7%; MSA, 86.7%; and PSP, 63.3%. Overall accuracy: 84.9%.

In line with this, adding the V5FC to the IVDr-6M-BB improved classification performance not only for HC (28/29 correctly classified; 96.6%; +10.3% absolute gain compared to the six-metabolites model), MSA (24/30; 80.0%; +3.3% absolute gain), but in particular for PSP (21/30; 70.0%; +10.0% absolute gain). However, the sensitivity was slightly reduced compared with the base IVDr-6M-BB (24/30 PD; 80.0%; −3.3% absolute loss) (Supplementary data, Figure S4 and Table S4). In contrast, adding both citrate and V5FC provided a near-perfect sensitivity (29/30 PD; 96.7%; +13.4% absolute gain compared to the six-metabolites model), and strong classification performance for HC (25/29; 93.1%; +6.9% absolute gain) and MSA (26/30; 86.7%; +10% absolute gain). PSP classification also improved modestly (19/30; 63.3%; +3.3% absolute gain) (Figure 5E). Importantly, here again, PSP scores from this extended biomarker correlated positively with disease duration since diagnosis, but also with UPDRS-III scores (Spearman’s ρ=+0.41 and +0.39, respectively, p < 0.05), further supporting an association between higher scores and more advanced disease (Supplementary data, Figure S2). For completeness, AD samples remained well classified (28/33; 84.8%; +6.0% absolute gain) (Supplementary data, Figure S5). All the classifications are summarized in Supplementary Figure S6.

Overall, augmenting the IVDr-6M-BB with V5FC and citrate substantially increased accuracy—94.9% for PD vs. HC (previously 84.7%), and 84.9% when including PSP/MSA (previously 76.5%)— representing +8-10% absolute gains across diagnosis distinctions.

## DISCUSSION

Although numerous previous studies have addressed the challenge of insufficient accuracy in diagnosing PD, there is still no fully validated reliable biomarker for its early and more specific detection (*23*), notably among different parkinsonian syndromes. This gap significantly hampers both clinical practice and translational research, limiting progress toward curative therapies and improving patient care. It also hinders the recruitment of homogeneous and well-characterized cohorts for clinical trials (*8*).

In the present study, we addressed this crucial clinical challenge by evaluating a blood biomarker, the 6M-BB, composed of a specific combination of six metabolites—acetoacetate, betaine, BHB, creatine, pyruvate, and valine, that we recently patented for PD discrimination from HC at the *de novo* stage (*41*). For that, we tested the 6M-BB using a wide range of new cases, from HC to *de novo* PD, as well as non-confounding neurodegenerative disorders such as AD, or confounding conditions, particularly atypical parkinsonian syndromes such as PSP and MSA. First, we confirmed the performance of the 6M-BB for discriminating PD patients from HC, with an accuracy of nearly 88%, demonstrating the reliability and robustness of the biomarker. We also showed its ability to properly exclude non-PD conditions. In particular, the majority of MSA and PSP patients were correctly classified as non-PD, despite their considerable and challenging overlap regarding clinical and pathological features with PD. Additionally, we successfully implemented this biomarker into the fully automated and standardized Bruker Avance IVDr NMR platform, representing a further step toward its application for PD diagnosis in clinical practice. Using this technology, we not only confirmed diagnosis performance comparable to the original 6M-BB, but also identified additional candidate metabolites and lipoproteins that can further enhance performance.

Despite numerous inconsistencies in the literature regarding PD-related metabolic dysregulations (*40, 42, 43*), we replicate in the present study most of those already described in our previous work (*41*). In particular, we consistently observed significant alterations in acetoacetate, betaine, BHB, and valine levels in 3 distinct *de novo* PD cohorts (n=21–30 each) from the USA and Italy, as well as in various PD animal models (*41*). In contrast, we observed a significant decrease in leucine in *de novo* PD patients whereas it was previously reported to be increased—an inconsistency often noted across studies (*42, 43*). We also report dysregulations in alanine, histidine, and lysine levels, in line with literature (*40, 42, 43*), while no significant changes were observed in our previous study (49). This could be linked to differences in sample preparation, pre-analytical variations in serum handling, or disease progression (*40, 42, 43*), but also to the impact of environment and lifestyle. Overall, this emphasizes the relevance of our strategy for selecting metabolites based not only on PD patients but also on complementary PD animal models (*44*). This also highlights the power of using combinations of metabolites instead of a single one as biomarker, as well as the crucial need for robust standardization methods in metabolomics applied to the clinic (*42*).

Consistently, the 6M-BB exhibits highly reproducible performance for diagnosis of early-stages PD, with a sensitivity approaching 87% and a specificity exceeding 89%, corresponding to an accuracy of nearly 88%. This level of performance was already achieved in our last publication (83% accuracy), with 73 drug-naive *de novo* PD and 53 HC (*41*). Altogether, these findings highlight the strong potential of this biomarker for clinical application. In parallel, many other candidate metabolomics biomarkers in biofluids have been published in recent years, showing similarly promising diagnostic performance (*45–47*). However, none of them combine such a rationale based on: i) high diversity of cohorts, ii) external validation on a large and independent cohort with demonstrated reproducibility, and iii) the exclusive recruitment of *de novo* PD patients who have never been treated with L-DOPA—a design still uncommon in biomarker research due to the difficulty of accessing untreated PD individuals.

In addition, we replicated these results on the IVDr 600 MHz NMR platform. It offers high degree of automation and standardization, making it particularly well-suited for routine clinical metabolomics and promising for the development of multicenter studies with robust inter-laboratory testing (*48*), facilitated by the growing number of IVDr NMR facilities. Numerous studies have already highlighted the strong potential of this technology for implementing NMR-based blood biomarkers in clinical practice, particularly in the fields of cardiometabolic syndromes (*49, 50*), cancer (*51, 52*), and SARS-CoV-2 infection (*53, 54*). However, only very few studies have focused on neurodegenerative diseases, particularly PD (*47, 55*). Among them, Meoni and colleagues reported that the overall serum metabolomic fingerprint, including both metabolites and lipoproteins, could achieve accurate discrimination between drug-naive *de novo* PD patients and HC with approximately 75% accuracy (*55*). In comparison, the 84.7% accuracy achieved here using the IVDr NMR platform appears highly promising. This persistent high performance is encouraging for the IVDr-6M-BB, with only a slight and not statistically significant decrease in accuracy in comparison with the 6M-BB.

Beyond distinguishing PD from HC, differential diagnosis between PD and other parkinsonian syndromes is also crucial for effective clinical management (*8*). Here, we report strong performance of our 6M-BB in distinguishing non-PD conditions, achieving approximately 88% accuracy for AD patients, and 76% and 72% for MSA and PSP, respectively, without auxiliary clinical information. Even if discriminating PD from AD is not clinically relevant, as they have clearly distinct symptoms in their early-phases (*56*), our results demonstrate that the biomarker reflects more than a general signature of neurodegeneration and is preferentially associated with PD pathogenesis. Furthermore, this study provides additional insight into PD and AD pathophysiological mechanisms. In particular, we show that, although AD and PD share several overlapping metabolic alterations, such as those involving creatine and valine, the directionality of their dysregulations is reversed compared to PD, with increased valine and decreased creatine levels in AD compared to healthy individuals. These differences were expected (*57, 58*) but still raise considerations that should be investigated more thoroughly. In contrast to AD, the differential diagnosis of PD against other atypical parkinsonian syndromes based on clinical evaluation is still currently highly challenging (*15, 17*). Previous studies estimated diagnostic accuracy to range from 62% to 79% for MSA (*59*) and approximately 80% for PSP (*60*), with both pathologies being among the most frequently diagnosed in case of false-positive PD (*61*). In addition, most biomarkers that correctly discriminate PD from atypical parkinsonian syndromes tend to perform much less well when it comes to distinguishing PD from HC (*29, 62*). By contrast, with an overall accuracy of ∼81% when considering PD, HC, but also MSA and PSP, the 6M-BB clearly outperforms many alternatives, both in distinguishing PD from HC and from atypical parkinsonian syndromes. Concerning the IVDr-6M-BB, its performance is slightly reduced with an overall accuracy of 76.5%, primarily due to a higher number of PSP patients being misclassified. This observation is consistent with the fewer metabolic alterations seen in PSP patients than in MSA relative to PD (*39, 63*). Nevertheless, Pathan and colleagues identified valine as an important discriminative variable for distinguishing PD from PSP (*39*). This may partly explain the reduced performance of the IVDr-6M-BB, as smaller valine differences are detected at 600 MHz than at 950MHz, a discrepancy that should be investigated further. Moreover, although acetoacetate and BHB had already been reported as key markers for distinguishing MSA from PD (50), we confirmed these findings and further demonstrated their relevance for distinguishing PSP from PD. Additionally, we found pyruvate to be among the top-10 variables for the distinction of PD against both MSA and PSP. Although pyruvate itself is not significantly altered in PD patients, its prior selection based on PD animal models (*41*), together with the known implication of altered glucose metabolism in the pathophysiology of PD (*39, 42, 64*), as well as its strong contribution to differentiating PD from atypical parkinsonian syndromes, underscores its crucial role in the diagnostic performance of the 6M-BB. Moreover, these results point to distinct metabolic pathway-level alterations between PD and MSA or PSP, notably a pronounced impairment of ketone body metabolism and branched-chain amino acid metabolism in PD, which is absent or only minimally observed in these two atypical parkinsonian syndromes (*39*).

A major added value of the Bruker IVDr NMR platform is the access to a broader range of metabolic variables, particularly a very large panel of lipoproteins, including particle concentrations, sizes, and detailed lipid content into HDL, LDL, and VLDL subclasses. Multiple studies report lipids and lipoproteins as key contributors to PD pathophysiology (*65–67*). Interestingly, we identified total HDL triglycerides (HDTG) as relevant variable for improving the sensitivity of the IVDr-6M-BB, while VLDL-5 and HDL-2 free cholesterol subclasses (V5FC and H2FC) enhanced its specificity. Alteration in HDL triglycerides have not previously been reported in PD, in contrast to LDL triglycerides (*55*). Although changes in triglyceride levels have been widely associated with PD (*68*), detailed profiling of lipoprotein subclasses in PD serum—such as HDL and LDL triglycerides—has been rarely reported (*47, 55*). Further studies involving larger and independent cohorts are therefore needed to draw definitive conclusions. With regard to cholesterol, many studies have highlighted an association between serum cholesterol levels over time and the risk of developing PD (*69, 70*). In the brain, evidence suggests that increased cholesterol and free cholesterol derived metabolites, such as oxysterols, contributes to the initiation of the αSyn aggregation and have a significant role in PD pathogenesis (*71, 72*). Concerning the HDL-2 free cholesterol (H2FC), Meoni and colleagues have previously reported dysregulation of total HDL free cholesterol in PD patients (*55*). However, here we found a non-significant decrease in PD as well as in MSA and PSP samples, limiting its diagnostic utility. Regarding VLDL-5 free cholesterol (V5FC), we observed an increase exclusive to PD and V5FC emerged as one of the most important variables for distinguishing PD from these two atypical parkinsonian syndromes. While serum VLDL cholesterol has previously been shown to be reduced in a cohort of PD patients spanning early to late stages (*73*), an increase was recently reported in sporadic PD patients, more pronounced in genetic PD (*47*). Moreover, in a prior study, prion inoculation has been shown to specifically increase the free cholesterol content of cultured cells (*74*), in line with αSyn aggregation. Regarding MSA and PSP, no data about free cholesterol are available to date. Altogether, the role of cholesterol—in particular of VLDL free cholesterol—should be further investigated in relation to the pathophysiology of these different parkinsonian syndromes. Nevertheless, V5FC already stands out as a promising addition to improve the IVDr-6M-BB for PD differential diagnosis.

Importantly, we also identified that citrate enhanced the sensitivity of the IVDr-6M-BB. Although we observed only a trend for higher citrate levels in PD patients, such alterations have already been reported in several studies, especially in late-stages PD patients (*47, 75*). Furthermore, a positive correlation between the L-DOPA equivalent daily dose (LEDD) and the citrate levels was observed in sporadic PD patients (*47*). Overall, these results suggest a relationship between citrate levels and PD progression. Moreover, it is known that the TCA cycle, which involves citrate, is altered in many neurodegenerative diseases including PD (*42, 58*). This emphasizes that citrate levels could reflect, at least in part, PD-related pathophysiological processes.

Finally, adding both V5FC and citrate to the IVDr-6M-BB increased the overall accuracy to almost 85%, with 93% of HC individuals and 97%, 87%, and 63% of PD, MSA and PSP patients, respectively, correctly classified. This clearly exceeds the accuracy currently achieved in clinical practice for differential diagnosis between PD and MSA (*10, 11, 59*), which appears to be more challenging than PSP (*60, 61*). Regarding PSP, the positive correlation between 6M-BB scores and disease duration, along with the shorter disease duration in correctly classified PSP cases, suggests that this biomarker is more effective at distinguishing early PSP cases from *de novo* PD than at advanced stages, where PSP is rarely mistaken for PD in clinical practice.

Altogether, our results demonstrate the robustness of our 6M-BBfor detecting PD at the *de novo* stage, as well as its relevance for differential diagnosis against atypical parkinsonian syndromes. Moreover, here we showed that IVDr-6M-BB, i.e. the 6M-BB transferred on the IVDr NMR platform, could help to bridge the gap between research and clinics. Lastly, we observed a highly promising improvement of the IVDr-6M-BB with the addition of citrate and V5FC. To our knowledge, this represents one of the most advanced metabolic biomarker approaches currently positioned for clinical differential diagnosis of PD. The speed of the entire diagnosis process, from sample preparation to classification—i.e. less than one hour regardless the number of molecules included in our metabolic biomarker—is a clear added value.

Nevertheless, it will be necessary to challenge this biomarker with a much bigger cohort of MSA and PSP patients, arising from different biobanks. Additionally, further efforts are needed to assess the consistency of our biomarker with disease progression and its correlation with clinical scales, and in particular to confirm the correlation highlighted here with the UPDRS-III score. This is highly relevant not only for clinicians, but also for deeper understanding of the relationship between phenotypic features of PD and underlying molecular alterations. It would also be particularly interesting to assess whether this biomarker can predict symptom fluctuations and/or disease worsening, two major patient-specific events that occur in the later clinical stages of PD. In addition, similar investigations will be crucial at earlier stages by evaluating whether our biomarker could predict the development of PD in prodromal individuals. This evaluation will help us to determine whether the prognostic value of the biomarker can reach the level of its differential diagnosis performance, and whether its differential diagnosis abilities hold when used at prodromal stages. Beyond the biomarker itself, deeper interrogation of the metabolite and lipoprotein dysregulations observed across the different parkinsonian syndromes could lead to a better understanding of the pathophysiological mechanisms associated with these diseases, and the identification of metabolic actors that could be investigated in more depth.

In conclusion, we confirmed the strong differential diagnostic performance of a blood biomarker of PD. Because PD, MSA, and PSP overlap clinically yet diverge in prognosis, treatment response, and trial eligibility, a serum-based, automated, and externally validated differential score has immediate clinical utility: it can streamline early triage, reduce inappropriate dopaminergic escalation in atypical parkinsonian syndromes, and enrich clinical trials with correctly classified participants. Moreover, the combined use of this biomarker with others reflecting other pathophysiological aspects of the disease could allow a more comprehensive biological classification of PD. This aligns with the goals of the SynNeurGe research diagnostic criteria (*76*), which proposes the first PD biological classification by combining biomarkers for pathological αSyn, PD-associated neurodegeneration, and pathogenic genetic variants. Our metabolic biomarker could therefore further enhance this type of classification. Together, these findings represent a step toward an improved biological classification of PD, bringing us closer to a truly effective diagnosis and patient stratification.

## MATERIALS AND METHODS

### Study design

This study aimed to evaluate the performance of a previously patented 6-metabolites blood biomarker (6M-BB) in differentiating PD from MSA and PSP, and to facilitate its translation to a clinical IVDr NMR platform. To this end, serum samples from *de novo* PD patients never treated with dopaminergic treatments (n=30), matched HC subjects (n=29), MSA patients (n=30), and PSP patients (n=30), were obtained from the Parkinson’s Disease Biomarkers Program (PDBP) consortium, supported by the National Institutes of Health (NIH) in USA. All these samples (n=119) were both analyzed by ^1^H-NMR at 950 MHz and 600 MHz. AD patients serum samples (n=50), were obtained from the TRYALZ database through the S-PARK clinical project (Grenoble Alpes University Hospital, CHUGA). These samples were provided in three successive batches: batches 1 (n=17) and 2 (n=16) were analyzed at 950 MHz (n=33 in total), and batches 2 and 3 (n=17) were analyzed at 600 MHz (n=33 in total). Patient inclusion criteria are described in Supplementary Materials. All samples were analyzed in a blinded manner and data exclusion criteria are described in Supplementary Materials.

### Sample collection

Blood was collected after at least 8 hours of fasting, or, if not feasible, after at least 8 hours of low-fat diet. They were centrifuged at 1500 g at 4°C for 10-15 minutes after coating at room temperature 15-60 min. Serum aliquots were stored at −80°C until further analyses.

### 950 MHz ^1^H-NMR analysis

Serum samples were submitted to the whole 950 MHz NMR metabolomics workflow as previously described in (*41*). Briefly, 3 mm NMR tubes were filled with 180 μL of diluted serum (60 μL serum and 120 μL PBS 0.1 M in 50% D_2_O, 7.4 pH). Experiments were performed at 300 K on a Bruker Advance III NMR spectrometer at 950 MHz (Infranalytics, IBS, Grenoble) using a Carr Purcell Meiboom Gill (CPMG) pulse sequence to attenuate macromolecule signals. The spectra were aligned on alanine peak using the Bruker software Topspin (version 4.1.1), then segmented using NMRProcFlow (version 1.4, online, http://nmrprocflow.org) in 0.001 ppm buckets between 0 and 10 ppm, with exclusion of residual water peak and ethanol signals. Each bucket was normalized to the total sum of buckets and the relative concentrations of metabolites were obtained as previously described in (*41*). All identified metabolites are listed in Supplementary data, Table S5.

### 600 MHz ^1^H-NMR analysis

Sample preparation and data acquisition followed Bruker IVDr B.I.Methods standard operating procedures (SOP) for blood plasma and serum (Bruker IVDr Methods User Manual, Bruker BioSpin GmbH & Co. KG, Germany), as previously described (*47, 50, 55*). Briefly, after thawing, 150 µL of serum were mixed with 150 µL of Bruker plasma buffer (Bruker BioSpin) and 3 mm 4” NMR SampleJet tubes (Bruker BioSpin) were manually filled with 200 μL of prepared serum. NMR spectra were acquired using a Bruker Avance IVDr platform (Avance Neo 600 MHz, Gemeli, IBP, Grenoble) equipped with a 5 mm BBI probe. As previously detailed (*50*), absolute quantification of 41 small molecules was performed using B.I.QUANT-PS2.0 package (Bruker IVDr Quantification of Plasma/Serum, Bruker IVDr software) and 112 lipoprotein classes and subclasses were quantified using the B.I.LISA package (Bruker IVDr Lipoprotein Subclass Analysis, Bruker IVDr software). As detailed in the Supplemental Materials, specific variables exclusion criteria were applied, resulting in a final inclusion of 26 metabolites (16 excluded) and 112 lipoprotein subfractions. Comprehensive information regarding all analyzed and excluded serum metabolites and lipoproteins is available in Supplementary Data, Table S6.

Betaine concentrations were estimated from two-dimensional J-resolved (2D-JRES) NMR datasets acquired according to the Bruker IVDr B.I.Methods standard operating procedures. Among the two characteristic NMR resonances of betaine, the methyl group signal (comprising nine equivalent protons) at 3.3 ppm was selected for primary analysis due to its higher signal to noise ratio. More details about the calibration and validation of betaine quantification can be found in Supplemental Materials.

## Statistical analyses

### Univariate statistics

Metabolomic data were analyzed in relation to the associated diagnosis only, without taking demographics (e.g., age, sex) into account, neither as explanatory variables nor as variables to be explained. All univariate analyses were performed using GraphPad PRISM (version 8.4.3). Results were expressed as mean ± SEM, with a 0.05 significance threshold. Normality was assed using Shapiro–Wilk tests. Given the non-normal distribution of some molecules at both 950 MHz and 600 MHz NMR, non-parametric tests were therefore uniformly applied: Mann–Whitney tests for pairwise comparisons, and Kruskal–Wallis tests followed by Dunn’s post hoc tests for multiple comparisons.

### Data compression and visualization

For a comprehensive visualization of all data, we used compression and projection methods, namely principal components analyses (PCA) and Orthogonal Partial Least Square–Discriminant Analysis (OPLS-DA), with the SIMCA software (Umetrics, version 17); detailed methodological parameters are provided in the Supplementary Materials.

### Classification and biomarker building

For classification and biomarker building, all analyses were performed using RStudio (version 4.4.2, packages used: *dplyr*, *glmnet*, and *pROC* packages).

#### Classification using the 6-metabolites blood biomarker (6M-BB)

The 6M-BB logistic regression—logit(P) = log(P/(1 − P)) = 42.79 + 0.17 BHB – 3.31 pyruvate – 12.01 valine + 7.06 acetoacetate + 3.14 creatine – 7.80 betaine (patent n° PCT/FR2021/052035)—was used to calculate the resulting score of each patient or control. The metrics used to evaluate the performance of the model for the classification of HC and PD cases were the area under the curve (AUC) of the receiver operating characteristic (ROC) curve (with a 95% confidence interval), the sensitivity, the specificity, and the accuracy (the number of correctly predicted cases divided by total cases) using a threshold of 0.9567. Contingency tables for all groups were generated based on the calculated scores and the same threshold.

#### Biomarker building with 600 MHz IVDr NMR data (IVDr-6M-BB)

new regression model, referred as IVDr-6M-BB, was build using PD and HC groups, as previously (*41*). The same metrics as for the 6M-BB were used to evaluate the performance of the model, calculated using a threshold of 0.51. A ROC curve obtained through leave-one-out cross-validation (LOO-CV) was also generated to check for potential overfitting. Contingency tables for all groups were generated based on the calculated scores

#### Increasing performance of the IVDr-6M-BB

selection of additional metabolites and lipoproteins was performed using feature selection and model optimization using Least Absolute Shrinkage and Selection Operator (LASSO) (α=1) with cross-validation, while forcing six core metabolites into the model based on prior biological knowledge. Candidate variables were further evaluated through individual logistic regression models and combinatorial approaches, with performance assessed via ROC analysis (AUC, sensitivity, specificity, PPV, NPV, accuracy) and LOO-CV validation. The relevance of each retained variable was assessed by comparing group means across PD, HC, and atypical parkinsonisms and by examining their importance in OPLS-DA models. Detailed statistical procedures and full model specifications are provided in Supplementary Materials.

## Supporting information

Graphical Abstract

## Data Availability

All data produced in the present study are available upon reasonable request to the authors

## Acknowledgments

Data and biospecimens used in preparation of this manuscript were obtained from the Parkinson’s Disease Biomarkers Program (PDBP) Consortium, supported by the National Institute of Neurological Disorders and Stroke at the National Institutes of Health. Investigators include: Roger Albin, Roy Alcalay, Alberto Ascherio, Thomas Beach, Sarah Berman, Bradley Boeve, F. DuBois Bowman, Shu Chen, Alice Chen-Plotkin, William Dauer, Ted Dawson, Paula Desplats, Richard Dewey, Ray Dorsey, Jori Fleisher, Kirk Frey, Douglas Galasko, James Galvin, Dwight German, Steven Gunzler, Lawrence Honig, Xuemei Huang, David Irwin, Un Kang, Kejal Kantarci, Anumantha Kanthasamy, Daniel Kaufer, Horacio Kaufmann, Qingzhong Kong, James Leverenz, Allan Levey, Carol Lippa, Irene Litvan, Oscar Lopez, Jian Ma, Richard Mailman, Lara Mangravite, Karen Marder, Kelly Mills, Nandakumar Narayanan, Laurie Orzelius, Vladislav Petyuk, Judith Potashkin, Liana Rosenthal, Rachel Saunders-Pullman, Clemens Scherzer, Michael Schwarzschild, Nicholas Seyfried, Tanya Simuni, Andrew Singleton, David Standaert, Debby Tsuang, David Vaillancourt, Jerrold Vitek, David Walt, Andrew West, Cyrus Zabetian, and Jing Zhang. The PDBP Investigators have not participated in reviewing the data analysis or content of the manuscript. The authors would like to thank Adrien Favier and Alicia Vallet from IBS (Grenoble) for technical assistance concerning 950 MHz NMR experiments and Infranalytics for financial support for conducting the research. They also thank the Gemeli platform (Grenoble) and in particular Joran Villaret for technical assistance concerning IVDr 600 MHz NMR experiments

## Funding

French National Research Agency grant ANR-24-CE17-3970-01 (SB, FF)

France Parkinson association grant R22185CC (SB, FF)

CerCoG LabEx grant C7H-LXCERCOG-GIN-1M25 (SB, FF)

Institut National de la Santé et de la Recherche Médicale - institutional funding

Grenoble Alpes University - institutional funding

## Author contributions

Conceptualization: VM, DM, SB, FF

Providing samples: MS, ALG.

Conducting experiments: VM, DM.

Acquiring data: VM, DM, SB, FF.

Processing data: VM, DM, CC, NP.

Analyzing data: VM, SB, FF, SC, EB

Funding acquisition: SB, FF

Supervision: SB, FF

Writing – original draft: VM, DM, SB, FF

Writing – review & editing: VM, DM, ALG, SB, FF

Co–first author order was determined by mutual discussion and agreement.

## Competing interests

The authors Claire Cannet and Nils Pompe are employees of Bruker BioSpin GmbH & Co. The authors Sabrina Boulet, Florence Fauvelle, and David Mallet are inventors of an awarded patent for the biomarker 6M-BB used in this study (patent n° PCT/FR2021/052035). A patent application for the extended IVDr biomarker has been filed listing Florence Fauvelle, Sabrina Boulet, David Mallet, and Vanille Millasseau as inventors (filing number: FR2602056).

## Data and materials availability

All raw data are available in the main text or the supplementary materials. Basic clinical metadata and Raw NMR quantitation data are available in the Excel file “*Raw Data*”. Supporting values for all figures are provided in the Excel file “*Supporting Data Values*.”

## Notes

### Author Declarations

All biological samples used in this study were obtained from established biobanks with their own ethical approvals and regulatory authorisations and were fully de-identified prior to use. Samples from the Parkinson's Disease Biomarkers Program (PDBP) were accessed through the PDBP biobank, coordinated by the National Institute of Neurological Disorders and Stroke (NINDS, National Institutes of Health, Bethesda, Maryland, USA). The PDBP is a multicenter initiative involving clinical sites across several U.S. states, including Texas, Florida, Pennsylvania, New York, Washington, Illinois, Maryland, and Massachusetts. Ethical approval for sample collection and utilisation in future research was obtained from the respective Institutional Review Boards (IRBs) of each participating site, affiliated with academic medical centres, such as the Institutional Review Board of Columbia University Irving Medical Centre (New York), for example. Samples used in the S-PARK project were obtained from the TRYALZ database (Grenoble University Hospital, France), whose ethical approval for sample collection and utilisation was given by the Delegation for Clinical Research and Innovation (Delegation a la Recherche Clinique et a l'Innovation, DRCI, France) In each case, ethical approval was granted prior to study initiation, and all participants provided written informed consent for participation and for the use of their biological samples in future research. All procedures performed in this study were conducted in accordance with the ethical standards of the Declaration of Helsinki (World Medical Association) and the Good Clinical Practice (GCP) guidelines.

