## Supplemental Data for "Toward clinical implementation of a metabolic blood biomarker for Parkinson’s disease differential diagnosis"

### Supplemental information

#### Supplemental Materials and Methods

##### Study design

*Inclusion criteria*. Inclusion criteria for PD patients included recent diagnosis (diagnosed ≤ 3 year) according to the Movement Disorders Society criteria (*77*), absence of dopaminergic treatments at the time of sampling, and absence of dementia or other medical conditions—particularly active psychiatric disorders—that could compromise the study. Inclusion criteria for controls were absence of neurological disease, family history of movement disorders and specific medical conditions. Inclusion criteria for MSA patients included established diagnosis according the Second Consensus Statement on the diagnosis of MSA (*78*), whereas PSP inclusion criteria included established diagnosis based on the International Parkinson's and MDS diagnostic criteria for PSP (*79*). Inclusion criteria for AD consisted of patients with minor to major cognitive impairments, with a AD confirmed by the AT(N) criteria and especially a CSF lumbar puncture A positive (Aß1-42, Aß1-40 and Aß1-42/Aß1-40 ratio) and T positive (Tau and p-Tau), relative to the 2018 NIA-AA guidelines for AD (*80*).

*Data exclusion criteria and outliers evaluation*. Data exclusion criteria were established prospectively and were based on $^{1}$H-NMR spectra quality regarding resolution and signal-to-noise ratio. In the 950 MHz $^{1}$H-NMR dataset, two spectra from the PDBP cohort (one HC and one MSA) were excluded due to excessive residual water signals. Dataset outliers were identified and evaluated using Grubbs' test.

##### 950 MHz $^{1}$H-NMR analysis

Serum samples were slowly thawed at 4°C. 3 mm NMR tubes were filled with 180 µL of diluted serum (60 µL serum and 120 µL PBS 0.1 M in 50% $D_2O$, 7.4 pH). $^{1}$H-NMR experiments were performed at 300 K on a Bruker Advance III NMR spectrometer at 950 MHz (Infranalytics, IBS, Grenoble), using a 5 mm cryoprobe One-dimension spectra were systematically recorded using a CPMG pulse sequence for edition of metabolite, with TE = 120 ms and 250 µs interpulse delay. The residual water signal was pre-saturated during 2 seconds of relaxation. Each spectrum using 128 averages lasted 10 minutes. After Fourier transformation, phasing and alignment on alanine peak using the Bruker software Topspin (version 4.1.1), more advanced preprocessing steps (baseline correction, local alignment and bucketing) were performed using NMRProcFlow (version 1.4, online, http://nmrprocflow.org). Assignment of peaks was performed using 2-dimension homonuclear $^{1}$H-$^{1}$H (TOCSY) and heteronuclear $^{1}$H-$^{13}$C (HSQC) spectra of selected samples, and database. When necessary,  addition of selected metabolites was used to unravel ambiguous assignments.

**600 MHz $^1$H-NMR analysis**

*Variable filtering:* several small-molecules were excluded from statistical analysis based on the following criteria: (i) high proportion of missing values (>30; (ii) low median sigCorr ($\rho < 75\%$) indicating low confidence in the quantification's reliability; and (iii) quality control variables (EDTA-related) or potential contaminant (ethanol), not relevant to the analysis. Lipoprotein variables were also filtered based on a high proportion of missing values; however, none of them met this exclusion criterion. To note, lipoproteins based on particle number (derived from Apo-B measurements) or as ratios were retained; nevertheless, caution was taken during data modeling to prevent bias from overlapping information. Finally, 26 metabolites (16 excluded) and 112 lipoproteins subfractions were included. All the information concerning the excluded and analyzed serum metabolites and lipoproteins are listed in Supplementary data, Table S6.

*Betaine quantification*. Calibration factors were established through a linear spiking series in pooled human serum samples, where known concentrations of betaine were incrementally added. The peak intensity of the methyl resonance in 2D-JRES spectrum was measured and correlated with the spiked concentrations to generate a calibration curve. This curve was subsequently applied to estimate betaine concentrations in study samples. Additionally, the CH2 group resonance of betaine was evaluated from the 2D JRES dataset as a secondary marker. However, this signal exhibited analytical interference with high glucose at concentrations >10 mM. To maintain consistency, estimated betaine concentrations from both methyl and methylene resonances were cross-checked, and samples showing significant inconsistencies, attributable to either significantly elevated glucose or TMAO concentrations, were flagged as outliers and excluded from the analysis. The methyl resonance of betaine at 3.3 ppm overlaps with trimethylamine N-oxide (TMAO) in human plasma and serum. While the described approach provides a robust estimate of betaine concentration, it is important to note that the method has not yet undergone fully analytical validation. Future work will include cross-validation against orthogonal techniques such as LC-MS to confirm accuracy and further investigate potential analytical interference. Until such validation is complete, the most accurate description of the evaluated resonance at 3.3 ppm is a composite signal representing both betaine and TMAO with betaine being the dominant contributor. Typical plasma concentrations of betaine and TMAO in large clinical cohorts were found to range between approximately 33-51 μM and 2.5-9.8 μM, respectively (*81*).

**Statistical Analysis**

*Data compression and visualization:* Unsupervised PCA allowed global visualization of the distribution of the samples, while supervised OPLS-DA allowed a better identification of important variables. For OPLS-DA, the total number of components was determined using the cross-validation procedure, which produces the R2Y and Q2 factors that indicate respectively the goodness of fit and the predictability of the model. A model is considered as robust and predictive when both factors were ≥ 0.5. Furthermore, analysis of variance of cross-validated predictive residuals (CV-ANOVA) was used to assess the significance of the model. Scores were plotted in 2D versus the two first principal components or versus first predictive/orthogonal components depending on the modelling. Loadings were mostly plotted in 2D. For the 950 MHz OPLS-DA models built with bucketed data, loadings were displayed in 1D to mimic a NMR spectrum (named "s-line" in SIMCA), in which peak direction indicates the sense of variation of corresponding metabolites, while color coding reflects their discrimination power. For the 600 MHz OPLS-DA models, VIP (Variable Importance in the Projection) scores were extracted and plotted using RStudio (version 4.4.2, ggplot2 package), ranked by their value, and colored based on the results of univariate statistics.

*Improvement of the IVDr-6M-BB.* Least Absolute Shrinkage and Selection Operator (LASSO) regularization was implemented using the *cv.glmnet* function, with the regularization parameter set to α=1. The six core metabolites were forced into the model based on prior biological knowledge by assigning them a penalty factor of 0, while the remaining metabolites were penalized using L1 regularization for automatic variable selection. All data were standardized prior to model fitting using z-scores. The regularization parameter (λ) was selected via 100-fold cross-validation by minimizing the binomial deviance. This approach shrinks some coefficients exactly to zero, enabling automatic variable selection. To further evaluate the individual contribution of each new selected variable, a series of logistic regression models were built, each including the core metabolites plus one of the LASSO-selected variables. This procedure was repeated separately for each candidate variable. Model performance was evaluated using ROC curve analyses. The following performance metrics were calculated: AUC with 95% confidence intervals, sensitivity, specificity, positive predictive value (PPV), negative predictive value (NPV), and accuracy. AUC was also estimated using LOO-CV. Combinatorial models including 2 to 3 of retained variables, added to the core metabolites, were then evaluated using the same ROC-based metrics.

**Study approval**

All biological samples used in this study were obtained from established biobanks with their own ethical approvals and regulatory authorisations. Samples from the Parkinson's Disease Biomarkers Program (PDBP) were accessed through the PDBP biobank, coordinated by the National Institute of Neurological Disorders and Stroke (NINDS, National Institutes of Health, Bethesda, Maryland, USA). The PDBP is a multicenter initiative involving clinical sites across several U.S. states, including Texas, Florida, Pennsylvania, New York, Washington, Illinois, Maryland, and Massachusetts. Ethical approval for sample collection and utilisation in future research was obtained from the respective Institutional Review Boards (IRBs) of each participating site, affiliated with academic medical centres, such as the Institutional Review Board of Columbia University Irving Medical Centre (New York), for example. Samples used in the S-PARK project were obtained from the TRYALZ database (Grenoble University Hospital, France) , whose ethical approval for sample collection and utilisation was given by the Delegation for Clinical Research and Innovation (Delegation à la Recherche Clinique et a l'Innovation, DRCI, France).

In each case, ethical approval was granted prior to study initiation, and all participants provided written informed consent for participation and for the use of their biological samples in future research. All procedures performed in this study were conducted in accordance with the ethical standards of the Declaration of Helsinki (World Medical Association) and the Good Clinical Practice (GCP) guidelines.

### Supplemental data

#### Supplemental tables

Supplemental Table S1

**Supplemental Table 1: Clinical characteristics of the cases**

| Patient | Cohort | Neurological examination | Diagnostic | Sex | Age range (ys) | Duration (mo) | H&Y | UPDRS I | UPDRS II | UPDRS III | Total UPDRS | MoCa | MMSE |
|---|---|---|---|---|---|---|---|---|---|---|---|---|---|
| **1** | PDBP | Case | MSA | F | 81-85 | 2 | 3 | 6 | 19 | 42 | 67 | 20 | - |
| **2** | PDBP | Case | MSA | F | 51-55 | 48 | 3 | 3 | 20 | 44 | 67 | 24 | - |
| **3** | PDBP | Case | MSA | M | 56-60 | 24 | 2 | 7 | 14 | 26 | 47 | 28 | - |
| **4** | PDBP | Case | MSA | M | 61-65 | 107 | 4 | - | - | 51 | - | 28 | - |
| **5** | PDBP | Case | MSA | F | 71-75 | 156 | 5 | 9 | 45 | 68 | 133 | 20 | - |
| **6** | PDBP | Case | MSA | M | 61-65 | 48 | 3 | 20 | 28 | 46 | 94 | 22 | - |
| **7** | PDBP | Case | MSA | M | 56-60 | 77 | 4 | - | - | 56 | - | 17 | - |
| **8** | PDBP | Case | MSA | M | 71-75 | 7 | 2 | 12 | 18 | 33 | 63 | 22 | - |
| **9** | PDBP | Case | MSA | F | 71-75 | 111 | 4 | 8 | 20 | 69 | 103 | 23 | - |
| **10** | PDBP | Case | MSA | F | 71-75 | 53 | 0 | - | - | 34 | - | 28 | - |
| **11** | PDBP | Case | MSA | M | 71-75 | 54 | 4 | - | - | 61 | - | 9 | - |
| **12** | PDBP | Case | MSA | M | 61-65 | 21 | 5 | 23 | 32 | 55 | 117 | 28 | - |
| **13** | PDBP | Case | MSA | F | 66-70 | 54 | 2 | - | - | 28 | - | 17 | - |
| **14** | PDBP | Case | MSA | M | 56-60 | 73 | 3 | - | - | 57 | - | 20 | - |
| **15** | PDBP | Case | MSA | M | 76-80 | 76 | 3 | 23 | 19 | 49 | 95 | 20 | - |
| **16** | PDBP | Case | MSA | M | 61-65 | 35 | 2 | - | - | 22 | - | 19 | - |
| **17** | PDBP | Case | MSA | M | 71-75 | 42 | 0 | - | - | 54 | - | 10 | - |
| **18** | PDBP | Case | MSA | M | 71-75 | 131 | 5 | 6 | 25 | 38 | 69 | 21 | - |
| **19** | PDBP | Case | MSA | M | 66-70 | 9 | 0 | - | - | 43 | - | 18 | - |
| **20** | PDBP | Case | MSA | M | 76-80 | 61 | 4 | - | - | 66 | - | 26 | - |
| **21** | PDBP | Case | MSA | F | 56-60 | 46 | 5 | 4 | 17 | 79 | 106 | 21 | - |
| **22** | PDBP | Case | MSA | M | 61-65 | 6 | 2 | 0 | 3 | 35 | 38 | 25 | - |
| **23** | PDBP | Case | MSA | M | 76-80 | 22 | 2 | 1 | 3 | 23 | 27 | 26 | - |
| **24** | PDBP | Case | MSA | F | 76-80 | 5 | 5 | - | - | 46 | - | 18 | - |
| **25** | PDBP | Case | MSA | F | 66-70 | 53 | 4 | - | - | 48 | - | 18 | - |
| **26** | PDBP | Case | MSA | F | 66-70 | 12 | 3 | - | - | 37 | - | 21 | - |
| **27** | PDBP | Case | MSA | M | 61-65 | 7 | 3 | - | - | 56 | - | 23 | - |
| **28** | PDBP | Case | MSA | F | 61-65 | 47 | 4 | 13 | 44 | 60 | 119 | - | - |
| **29** | PDBP | Case | MSA | M | 61-65 | 1 | 3 | 6 | 7 | 26 | 39 | 23 | - |
| **30** | PDBP | Case | MSA | M | 61-65 | 4 | 4 | - | - | 80 | - | 24 | - |
| **31** | PDBP | Control | HC | F | 56-60 | N/A | 0 | 0 | 0 | 0 | 0 | 27 | - |
| **32** | PDBP | Control | HC | F | 56-60 | N/A | 0 | 11 | 0 | 0 | 11 | 28 | - |
| **33** | PDBP | Control | HC | F | 56-60 | N/A | 0 | 8 | 1 | 0 | 9 | 29 | - |
| **34** | PDBP | Control | HC | F | 61-65 | N/A | 0 | 5 | 0 | 6 | 11 | 29 | - |
| **35** | PDBP | Control | HC | M | 56-60 | N/A | 0 | 0 | 0 | 0 | 0 | 23 | - |
| **36** | PDBP | Control | HC | F | 61-65 | N/A | 0 | 4 | 0 | 0 | 4 | 30 | - |
| **37** | PDBP | Control | HC | F | 66-70 | N/A | 0 | 3 | 0 | 1 | 4 | 25 | - |
| **38** | PDBP | Control | HC | M | 51-55 | N/A | 0 | 1 | 0 | 0 | 1 | 29 | - |
| **39** | PDBP | Control | HC | F | 66-70 | N/A | 0 | 0 | 0 | 2 | 2 | 23 | - |
| **40** | PDBP | Control | HC | F | 66-70 | N/A | 0 | 3 | 0 | 1 | 4 | 23 | - |
| **41** | PDBP | Control | HC | M | 66-70 | N/A | 0 | 6 | 3 | 0 | 9 | 27 | - |
| **42** | PDBP | Control | HC | M | 61-65 | N/A | 0 | 0 | 0 | 13 | 13 | 26 | - |
| **43** | PDBP | Control | HC | F | 56-60 | N/A | 0 | 3 | 2 | 0 | 5 | 29 | - |
| **44** | PDBP | Control | HC | M | 56-60 | N/A | 0 | 0 | 0 | 1 | 1 | 30 | - |
| **45** | PDBP | Control | HC | F | 61-65 | N/A | 0 | 2 | 0 | 0 | 2 | 27 | - |
| **46** | PDBP | Control | HC | F | 51-55 | N/A | 0 | 0 | 0 | 3 | 3 | 27 | - |
| **47** | PDBP | Control | HC | F | 61-65 | N/A | 0 | 6 | 0 | 0 | 6 | 26 | - |
| **48** | PDBP | Control | HC | F | 71-75 | N/A | 0 | 6 | 1 | 1 | 8 | 22 | - |
| **49** | PDBP | Control | HC | F | 51-55 | N/A | 0 | 3 | 0 | 1 | 4 | 29 | - |
| **50** | PDBP | Control | HC | F | 71-75 | N/A | 0 | 3 | 0 | 3 | 6 | 30 | - |
| **51** | PDBP | Control | HC | F | 56-60 | N/A | 0 | 0 | 0 | 1 | 1 | 30 | - |
| **52** | PDBP | Control | HC | F | 56-60 | N/A | 0 | 2 | 0 | 1 | 3 | 29 | - |
| **53** | PDBP | Control | HC | M | 66-70 | N/A | 0 | - | - | 2 | - | 28 | - |
| **54** | PDBP | Control | HC | M | 56-60 | N/A | 0 | - | - | 1 | - | 28 | - |
| **55** | PDBP | Control | HC | M | 66-70 | N/A | 0 | 2 | 0 | 3 | 5 | 29 | - |
| **56** | PDBP | Control | HC | M | 61-65 | N/A | 0 | 1 | 0 | 0 | 1 | 28 | - |
| **57** | PDBP | Control | HC | F | 61-65 | N/A | 0 | 6 | 1 | 0 | 7 | 28 | - |
| **58** | PDBP | Control | HC | M | 66-70 | N/A | 0 | 3 | 0 | 4 | 7 | 26 | - |
| **59** | PDBP | Control | HC | M | 76-80 | N/A | 0 | 4 | 2 | 2 | 8 | 28 | - |
| **60** | PDBP | Case | PD | M | 66-70 | 10 | 1 | 9 | 5 | 18 | 32 | 25 | - |
| **61** | PDBP | Case | PD | F | 66-70 | 22 | 2 | 19 | 8 | 19 | 51 | 27 | - |
| **62** | PDBP | Case | PD | M | 66-70 | -2 | 2 | 5 | 4 | 14 | 23 | 26 | - |
| **63** | PDBP | Case | PD | M | 61-65 | 8 | 1 | 10 | 3 | 6 | 19 | 30 | - |
| **64** | PDBP | Case | PD | M | 56-60 | 36 | 1 | 4 | 4 | 11 | 19 | 28 | - |
| **65** | PDBP | Case | PD | M | 66-70 | 8 | 2 | 19 | 13 | 19 | 51 | 26 | - |
| **66** | PDBP | Case | PD | M | 61-65 | <36 | 2 | 15 | 11 | 20 | 46 | 25 | - |

| | | | | | | | | | | | | | |
|---|---|---|---|---|---|---|---|---|---|---|---|---|---|
| **67** | PDBP | Case | PD | F | 56-60 | -1 | 0 | 8 | 6 | 4 | 18 | 23 | - |
| **68** | PDBP | Case | PD | M | 56-60 | <36 | 2 | 5 | 15 | 13 | 33 | 28 | - |
| **69** | PDBP | Case | PD | M | 66-70 | 8 | 1 | 4 | 5 | 11 | 20 | 28 | - |
| **70** | PDBP | Case | PD | F | 51-55 | <36 | 2 | 4 | 2 | 14 | 20 | 23 | - |
| **71** | PDBP | Case | PD | F | 61-65 | <36 | 2 | 7 | 4 | 38 | 49 | 26 | - |
| **72** | PDBP | Case | PD | F | 51-55 | 5 | 2 | 8 | 8 | 17 | 33 | 26 | - |
| **73** | PDBP | Case | PD | F | 56-60 | <36 | 2 | 0 | 3 | 7 | 10 | 28 | - |
| **74** | PDBP | Case | PD | F | 56-60 | <36 | 2 | 0 | 2 | 11 | 14 | 30 | - |
| **75** | PDBP | Case | PD | F | 81-85 | <36 | 2 | 23 | 19 | 15 | 59 | 27 | - |
| **76** | PDBP | Case | PD | M | 61-65 | <36 | 1 | 2 | 11 | 14 | 27 | 30 | - |
| **77** | PDBP | Case | PD | F | 56-60 | <36 | 2 | 14 | 6 | 10 | 30 | 30 | - |
| **78** | PDBP | Case | PD | M | 66-70 | 21 | 2 | 14 | 6 | 14 | 34 | 26 | - |
| **79** | PDBP | Case | PD | F | 51-55 | <36 | 2 | 6 | 11 | 16 | 33 | 26 | - |
| **80** | PDBP | Case | PD | M | 61-65 | 24 | 1 | 3 | 7 | 12 | 22 | 28 | - |
| **81** | PDBP | Case | PD | F | 56-60 | 36 | 1 | 3 | 8 | 18 | 29 | 29 | - |
| **82** | PDBP | Case | PD | M | 66-70 | 7 | 2 | - | - | 17 | - | 26 | - |
| **83** | PDBP | Case | PD | M | 66-70 | 0 | 1 | 3 | 3 | 4 | 10 | 25 | - |
| **84** | PDBP | Case | PD | F | 51-55 | 8 | 2 | 5 | 8 | 10 | 23 | 27 | - |
| **85** | PDBP | Case | PD | M | 61-65 | 33 | 2 | 7 | 8 | 40 | 55 | 27 | - |
| **86** | PDBP | Case | PD | M | 31-35 | <36 | 2 | 2 | 1 | 3 | 7 | 29 | - |
| **87** | PDBP | Case | PD | M | 61-65 | 3 | 1 | 2 | 4 | 10 | 16 | 24 | - |
| **88** | PDBP | Case | PD | F | 76-80 | 22 | 3 | 4 | 8 | 53 | 69 | 26 | - |
| **89** | PDBP | Case | PD | F | 66-70 | 9 | 1 | 2 | 1 | 19 | 22 | 24 | - |
| **90** | PDBP | Case | PSP | M | 61-65 | 1 | 2 | 6 | 22 | - | - | 29 | - |
| **91** | PDBP | Case | PSP | M | 56-60 | 0 | 3 | 7 | 21 | 44 | 72 | 26 | - |
| **92** | PDBP | Case | PSP | F | 66-70 | 33 | 3 | - | - | 37 | - | 18 | - |
| **93** | PDBP | Case | PSP | F | 66-70 | 28 | 0 | - | - | 33 | - | 25 | - |
| **94** | PDBP | Case | PSP | M | 66-70 | 3 | 2 | 2 | 12 | 28 | 42 | 23 | - |
| **95** | PDBP | Case | PSP | M | 71-75 | 9 | 0 | - | - | 30 | - | 24 | - |
| **96** | PDBP | Case | PSP | M | 71-75 | 0 | 2 | 4 | 10 | 28 | 42 | 29 | - |
| **97** | PDBP | Case | PSP | F | 81-85 | 6 | 3 | - | - | 35 | - | 24 | - |
| **98** | PDBP | Case | PSP | F | 61-65 | 10 | 5 | - | - | 67 | - | 15 | - |
| **99** | PDBP | Case | PSP | M | 61-65 | 44 | 5 | 12 | 43 | 54 | 109 | - | - |
| **100** | PDBP | Case | PSP | M | 66-70 | 109 | 2 | - | - | 18 | - | 27 | - |
| **101** | PDBP | Case | PSP | F | 66-70 | 24 | 4 | 12 | 26 | 65 | 104 | 19 | - |
| **102** | PDBP | Case | PSP | M | 61-65 | 29 | 4 | 29 | 51 | 98 | 199 | - | - |
| **103** | PDBP | Case | PSP | F | 66-70 | 0 | 2 | 13 | 10 | 43 | 66 | 22 | - |
| **104** | PDBP | Case | PSP | M | 66-70 | 40 | 4 | 12 | 40 | 78 | 141 | - | - |
| **105** | PDBP | Case | PSP | M | 71-75 | 8 | 3 | 5 | 17 | 46 | 71 | 24 | - |
| **106** | PDBP | Case | PSP | F | 71-75 | 55 | 4 | 13 | 17 | 55 | 94 | 29 | - |
| **107** | PDBP | Case | PSP | M | 76-80 | 13 | 2 | 11 | 20 | 21 | 52 | 18 | - |
| **108** | PDBP | Case | PSP | M | 61-65 | 98 | 3 | 3 | 40 | 55 | 98 | 20 | - |
| **109** | PDBP | Case | PSP | F | 71-75 | 12 | 5 | 15 | 44 | 54 | 114 | 12 | - |
| **110** | PDBP | Case | PSP | F | 66-70 | 2 | 3 | 5 | 19 | 43 | 67 | 21 | - |
| **111** | PDBP | Case | PSP | M | 71-75 | 27 | 0 | - | - | 61 | - | 17 | - |
| **112** | PDBP | Case | PSP | M | 56-60 | 23 | 3 | - | - | 50 | - | 27 | - |
| **113** | PDBP | Case | PSP | M | 61-65 | 18 | 2 | - | - | 53 | - | 26 | - |
| **114** | PDBP | Case | PSP | F | 61-65 | 6 | 5 | 5 | 27 | 21 | 53 | 25 | - |
| **115** | PDBP | Case | PSP | F | 71-75 | 66 | 0 | - | - | 60 | - | 19 | - |
| **116** | PDBP | Case | PSP | F | 71-75 | 58 | 0 | - | - | 52 | - | 11 | - |
| **117** | PDBP | Case | PSP | M | 66-70 | 46 | 0 | - | - | 40 | - | 23 | - |
| **118** | PDBP | Case | PSP | F | 56-60 | 23 | 0 | - | - | 59 | - | 23 | - |
| **119** | PDBP | Case | PSP | F | 66-70 | 28 | 4 | - | - | 61 | - | 25 | - |
| **120** | S-PARK | Case | AD | M | 71-75 | -1 | - | - | - | - | - | - | 25 |
| **121** | S-PARK | Case | AD | M | 66-70 | -3 | - | - | - | - | - | - | 16 |
| **122** | S-PARK | Case | AD | F | 76-80 | -2 | - | - | - | - | - | - | 20 |
| **123** | S-PARK | Case | AD | M | 76-80 | -3 | - | - | - | - | - | - | 26 |
| **124** | S-PARK | Case | AD | M | 66-70 | -2 | - | - | - | - | - | - | 10 |
| **125** | S-PARK | Case | AD | F | 76-80 | -3 | - | - | - | - | - | - | 22 |
| **126** | S-PARK | Case | AD | F | 56-60 | 2 | - | - | - | - | - | - | 23 |
| **127** | S-PARK | Case | AD | M | 76-80 | -2 | - | - | - | - | - | - | 22 |
| **128** | S-PARK | Case | AD | F | 71-75 | -2 | - | - | - | - | - | - | 19 |
| **129** | S-PARK | Case | AD | M | 81-85 | -3 | - | - | - | - | - | - | 28 |
| **130** | S-PARK | Case | AD | F | 81-85 | -2 | - | - | - | - | - | - | 14 |
| **131** | S-PARK | Case | AD | F | 71-75 | -3 | - | - | - | - | - | - | 20 |
| **132** | S-PARK | Case | AD | F | 61-65 | 2 | - | - | - | - | - | - | 11 |
| **133** | S-PARK | Case | AD | F | 66-70 | -3 | - | - | - | - | - | - | 18 |
| **134** | S-PARK | Case | AD | F | 66-70 | -2 | - | - | - | - | - | - | 10 |
| **135** | S-PARK | Case | AD | M | 66-70 | -1 | - | - | - | - | - | - | 24 |
| **136** | S-PARK | Case | AD | F | 51-55 | -2 | - | - | - | - | - | - | 22 |
| **137** | S-PARK | Case | AD | F | 61-65 | -2 | - | - | - | - | - | - | 14 |
| **138** | S-PARK | Case | AD | M | 71-75 | -2 | - | - | - | - | - | - | 21 |
| **139** | S-PARK | Case | AD | F | 61-65 | -2 | - | - | - | - | - | - | 17 |
| **140** | S-PARK | Case | AD | F | 71-75 | -3 | - | - | - | - | - | - | 25 |
| **141** | S-PARK | Case | AD | F | 71-75 | -2 | - | - | - | - | - | - | 13 |
| **142** | S-PARK | Case | AD | F | 76-80 | -2 | - | - | - | - | - | - | 19 |
| **143** | S-PARK | Case | AD | M | 76-80 | -2 | - | - | - | - | - | - | 18 |
| **144** | S-PARK | Case | AD | F | 71-75 | -2 | - | - | - | - | - | - | 26 |
| **145** | S-PARK | Case | AD | M | 61-65 | 1 | - | - | - | - | - | - | 16 |
| **146** | S-PARK | Case | AD | M | 61-65 | -3 | - | - | - | - | - | - | 19 |
| **147** | S-PARK | Case | AD | M | 66-70 | -2 | - | - | - | - | - | - | 27 |
| **148** | S-PARK | Case | AD | M | 71-75 | -2 | - | - | - | - | - | - | 16 |

| | | | | | | | | | | | | | |
|---|---|---|---|---|---|---|---|---|---|---|---|---|---|
| **149** | S-PARK | Case | AD | F | 61-65 | - | - | - | - | - | - | - | 10 |
| **150** | S-PARK | Case | AD | M | 66-70 | -1 | - | - | - | - | - | - | 9 |
| **151** | S-PARK | Case | AD | M | 76-80 | -3 | - | - | - | - | - | - | 23 |
| **152** | S-PARK | Case | AD | M | 76-80 | 0 | - | - | - | - | - | - | 23 |
| **153** | S-PARK | Case | AD | F | 76-80 | -2 | - | - | - | - | - | - | 17 |
| **154** | S-PARK | Case | AD | F | 76-80 | -10 | - | - | - | - | - | - | 15 |
| **155** | S-PARK | Case | AD | M | 76-80 | -2 | - | - | - | - | - | - | 24 |
| **156** | S-PARK | Case | AD | F | 76-80 | -2 | - | - | - | - | - | - | 21 |
| **157** | S-PARK | Case | AD | M | 66-70 | -3 | - | - | - | - | - | - | 19 |
| **158** | S-PARK | Case | AD | M | 66-70 | -2 | - | - | - | - | - | - | 23 |
| **159** | S-PARK | Case | AD | M | 71-75 | -2 | - | - | - | - | - | - | 19 |
| **160** | S-PARK | Case | AD | F | 76-80 | -1 | - | - | - | - | - | - | 20 |
| **161** | S-PARK | Case | AD | F | 61-65 | -2 | - | - | - | - | - | - | 22 |
| **162** | S-PARK | Case | AD | F | 71-75 | -3 | - | - | - | - | - | - | 19 |
| **163** | S-PARK | Case | AD | F | 76-80 | -3 | - | - | - | - | - | - | 15 |
| **164** | S-PARK | Case | AD | F | 71-75 | - | - | - | - | - | - | - | 20 |
| **165** | S-PARK | Case | AD | M | 76-80 | -2 | - | - | - | - | - | - | 14 |
| **166** | S-PARK | Case | AD | M | 76-80 | -3 | - | - | - | - | - | - | 22 |
| **167** | S-PARK | Case | AD | F | 61-65 | -2 | - | - | - | - | - | - | 19 |
| **168** | S-PARK | Case | AD | F | 51-55 | -3 | - | - | - | - | - | - | 20 |
| **169** | S-PARK | Case | AD | M | 66-70 | -2 | - | - | - | - | - | - | 24 |

A negative value for the duration since diagnosis corresponds to a blood sample taken before the diagnosis was confirmed . *F: Female; M: Male; yr: years; mo: months; Duration: Duration between neurological diagnosis and blood sampling, negative values indicate that blood sampling was performed before the diagnosis; H&Y: Hoehn and Yahr scale; UPDRS: Unified Parkinson disease rating scale; MoCA: Montreal Cognitive Assessment; MMSE: Mini-Mental State Examination; N/A = not applicable*.

Supplemental Table S2

**Table S2: Mean metabolite relative concentrations in serum analysed with 950 MHz NMR with standard deviations (SD) and Mann-Whitney test results with pairwise comparisons against PD group.**

| Metabolite | Mean (SD) in a.u. | | | | | *p* values (compared to PD) | | | |
|---|---|---|---|---|---|---|---|---|---|
| | PD (n=30) | HC (n=28) | MSA (n=29) | PSP (n=29) | AD (n=33) | HC | MSA | PSP | AD |
| **Formate** | 10.24 (5.71) | 9.71 (5.12) | 9.32 (5.38) | 10.38 (7.1) | 24.54 (5.54) | 6,80E-01 | 3,03E-01 | 2,17E-01 | **4,35E-10** |
| **Phenylalanine** | 52.8 (11.04) | 54.47 (11.72) | 53.24 (7.99) | 53.48 (10.4) | 96.47 (28.64) | 6,13E-01 | 7,05E-01 | 6,99E-01 | **1,04E-09** |
| **Histidine** | 58.09 (9.68) | 65.16 (9.21) | 61.1 (11.26) | 59.66 (13.7) | 88.17 (12.75) | **1,62E-02** | 1,87E-01 | 5,10E-01 | **1,25E-10** |
| **Tyrosine** | 102.48 (20.01) | 116.65 (31.63) | 112.79 (28.51) | 117.03 (27.53) | 121.98 (20.83) | 8,00E-02 | 2,19E-01 | **4,96E-02** | **1,03E-03** |
| **Proline** | 56.62 (27.07) | 49.93 (15.53) | 58.92 (29.13) | 62.37 (33.75) | 88.25 (34.1) | 5,81E-01 | 3,63E-01 | 2,72E-01 | **7,60E-05** |
| **Lactate** | 954.47 (437.38) | 804.27 (313.25) | 1035.28 (346.47) | 1023.77 (352.97) | 1662.22 (455.87) | 1,14E-01 | 2,49E-01 | 2,72E-01 | **7,14E-08** |
| **Ascorbate** | 87.25 (63) | 75.07 (34.06) | 95.12 (74.53) | 124.63 (93.75) | 78.01 (84.64) | 9,07E-01 | 3,63E-01 | **3,32E-02** | **1,83E-02** |
| **Serine** | 117.72 (34.54) | 107.11 (18.77) | 120.74 (34.62) | 126.81 (49.19) | 181.95 (44.51) | 2,16E-01 | 7,73E-01 | 8,02E-01 | **2,05E-08** |
| **Threonine** | 244.17 (49.94) | 242.36 (37.37) | 245.86 (55.96) | 264.41 (66.95) | 328.26 (47.9) | 7,26E-01 | 8,68E-01 | 2,34E-01 | **3,85E-08** |
| **Glycine** | 660.2 (160.33) | 614.46 (126.51) | 656.83 (201.94) | 682.61 (318.11) | 942.83 (144.3) | 2,73E-01 | 6,38E-01 | 5,80E-01 | **6,12E-08** |
| **Myo-inositol** | 70.1 (23.08) | 79.44 (18.62) | 73.12 (19.78) | 79.11 (23.06) | 105.4 (25.01) | 6,99E-02 | 4,22E-01 | 8,26E-02 | **1,41E-06** |
| **Betaine** | 369 (105.62) | 456.26 (122.25) | 393.74 (89.12) | 428.09 (206.82) | 471.76 (130.8) | **1,15E-02** | 3,47E-01 | 2,78E-01 | **1,66E-03** |
| **Choline** | 264.46 (54.24) | 249.76 (37.8) | 246.26 (47.63) | 240.08 (45.5) | 325.57 (67.76) | 2,34E-01 | 1,37E-01 | 9,39E-02 | **3,95E-04** |
| **DMSO2** | 46.77 (15.83) | 214.77 (870.29) | 397.71 (1288.99) | 45.79 (20.44) | 62.83 (31.94) | 8,34E-01 | 2,82E-01 | 5,19E-01 | 7,70E-02 |
| **Creatinine** | 160.69 (31.77) | 173.88 (25.42) | 178.81 (36.17) | 170.04 (34.98) | 186.67 (35.56) | 8,00E-02 | 1,08E-01 | 2,28E-01 | **6,30E-03** |
| **Creatine** | 110.73 (48.95) | 90.23 (34.21) | 94.44 (31.2) | 99.49 (37.4) | 96.55 (37.11) | 9,14E-02 | 2,37E-01 | 4,53E-01 | 2,92E-01 |
| **Asparagine** | 43.69 (9.92) | 45.33 (6.85) | 47.16 (12.62) | 43.59 (12.92) | 49.98 (9.02) | 3,79E-01 | 2,89E-01 | 6,44E-01 | **6,85E-03** |
| **N-dimethylglycine** | 24.54 (5.42) | 26.28 (7.94) | 24.61 (6.43) | 22.89 (5.73) | 28.24 (6.51) | 4,89E-01 | 9,76E-01 | 2,92E-01 | **1,76E-02** |
| **Methionine** | 82.84 (14.75) | 82.26 (11.06) | 81.78 (15.63) | 82.6 (16.28) | 88.71 (15.16) | 6,46E-01 | 8,32E-01 | 9,94E-01 | 5,85E-02 |
| **Citrate** | 249.13 (61.09) | 233.38 (50.89) | 251.88 (58.11) | 245.63 (59.56) | 260.58 (64.98) | 2,16E-01 | 7,05E-01 | 9,09E-01 | 6,06E-01 |
| **Glutamine** | 817.67 (189.44) | 837.88 (107.91) | 843.87 (167.76) | 852.62 (197.78) | 680.27 (150.23) | 5,81E-01 | 4,76E-01 | 5,90E-01 | **2,64E-03** |
| **Pyruvate** | 98.68 (37.51) | 103.54 (34.91) | 120.73 (47.11) | 139.21 (49.36) | 50.82 (14.37) | 4,60E-01 | **4,22E-02** | **3,77E-04** | **2,44E-09** |
| **Acetoacetate** | 124.19 (55.97) | 101.94 (35.51) | 90.38 (28.59) | 105.52 (45.62) | 77.63 (14.83) | 1,55E-01 | **2,29E-02** | 1,85E-01 | **5,06E-05** |
| **Acetate** | 284.28 (54.66) | 290.38 (53.36) | 275.16 (50.56) | 282.57 (61.85) | 422.13 (67.95) | 5,39E-01 | 7,50E-01 | 9,46E-01 | **1,34E-09** |
| **Lysine** | 589.71 (84.77) | 652.26 (87.61) | 642.62 (116.73) | 646.61 (150.65) | 856.99 (134.5) | **1,31E-02** | **3,78E-02** | 1,24E-01 | **2,06E-09** |
| **Arginine** | 84.34 (18.67) | 97.07 (24.28) | 99.54 (26.9) | 99.2 (25.06) | 114.99 (34.29) | 5,66E-02 | **2,39E-02** | **1,99E-02** | **1,50E-04** |
| **Alanine** | 1117.32 (184.77) | 1264.21 (223.24) | 1325.76 (283.47) | 1451.91 (398.79) | 1585.05 (326.49) | **7,27E-03** | **2,31E-03** | **5,02E-04** | **1,74E-09** |
| **BHB** | 273.88 (104.27) | 208.88 (62.96) | 204.67 (89.23) | 203.14 (66.26) | 229.18 (113.75) | **7,99E-04** | **2,03E-04** | **2,36E-04** | 1,00E-01 |
| **Valine** | 610.38 (101.52) | 710.88 (135.41) | 673.67 (122.39) | 698.14 (197.47) | 762.8 (110.76) | **1,05E-03** | **1,24E-02** | 5,91E-02 | **4,16E-06** |
| **Isoleucine** | 193.04 (44.08) | 215.71 (54.84) | 218.65 (50.79) | 230.29 (82.18) | 250.79 (50.16) | 8,27E-02 | **3,38E-02** | **4,45E-02** | **8,51E-06** |
| **Leucine** | 216.74 (38.55) | 241.85 (41.86) | 225.22 (43.43) | 236.4 (67.45) | 311.27 (46.23) | **4,74E-03** | 1,63E-01 | 3,75E-01 | **8,45E-09** |
| **Glucose** | 2485.04 (336.08) | 2640.43 (408.78) | 2629.61 (435.19) | 2676.51 (375.16) | 2563.35 (463.57) | 1,07E-01 | 1,50E-01 | 7,00E-02 | 6,06E-01 |

Significant p values of less than 0.05 are indicated in bold. *a.u.: arbitrary unit.*

Supplemental Table S3

**Table S3: Mean metabolite concentrations in serum analysed with 600 MHz NMR with standard deviations (SD) and Mann-Whitney test results with pairwise comparisons against PD group.**

| Metabolite | Mean (SD) in mmol/L | | | | | *p* values (compared to PD) | | | |
|---|---|---|---|---|---|---|---|---|---|
| | PD (n=30) | HC (n=28) | MSA (n=29) | PSP (n=29) | AD (n=33) | HC | MSA | PSP | AD |
| Betaine | 0,045 (0,018) | 0,063 (0,032) | 0,051 (0,019) | 0,058 (0,043) | 0,049 (0,016) | **3,38E-02** | 3,08E-01 | 2,74E-01 | 6,45E-01 |
| Trimethylamine-N-oxide | 0,022 (0,013) | 0,028 (0,022) | 0,02 (0,013) | 0,024 (0,023) | 0,02 (0,014) | 4,90E-01 | 7,28E-01 | 9,82E-01 | 5,00E-01 |
| Alanine | 0,397 (0,111) | 0,431 (0,073) | 0,465 (0,094) | 0,485 (0,126) | 0,562 (0,14) | **4,87E-02** | **3,67E-03** | **7,96E-03** | **6,29E-07** |
| Creatine | 0,030 (0,020) | 0,019 (0,013) | 0,022 (0,016) | 0,03 (0,024) | 0,022 (0,018) | **2,43E-02** | 6,88E-02 | 8,42E-01 | 5,57E-02 |
| Creatinine | 0,084 (0,022) | 0,084 (0,023) | 0,088 (0,02) | 0,08 (0,019) | 0,112 (0,026) | 8,79E-01 | 5,10E-01 | 4,96E-01 | **5,84E-05** |
| Glutamine | 0,724 (0,109) | 0,748 (0,068) | 0,758 (0,07) | 0,716 (0,089) | 0,493 (0,159) | 1,72E-01 | 7,72E-02 | 8,02E-01 | **2,05E-08** |
| Glycine | 0,335 (0,066) | 0,299 (0,056) | 0,323 (0,082) | 0,328 (0,136) | 0,38 (0,095) | **3,38E-02** | 3,22E-01 | 1,78E-01 | 8,29E-02 |
| Histidine | 0,076 (0,035) | 0,08 (0,029) | 0,082 (0,037) | 0,069 (0,025) | 0,127 (0,032) | 5,44E-01 | 4,55E-01 | 4,82E-01 | **2,91E-08** |
| Isoleucine | 0,052 (0,02) | 0,059 (0,018) | 0,057 (0,018) | 0,057 (0,025) | 0,061 (0,011) | 7,84E-02 | 2,20E-01 | 4,33E-01 | **1,07E-03** |
| Leucine | 0,096 (0,033) | 0,116 (0,036) | 0,103 (0,024) | 0,109 (0,049) | 0,127 (0,021) | **1,13E-02** | 1,35E-01 | 4,20E-01 | **8,46E-06** |
| Methionine | 0,038 (0,02) | 0,038 (0,021) | 0,035 (0,02) | 0,034 (0,021) | 0,07 (0,021) | 8,91E-01 | 6,52E-01 | 4,12E-01 | **8,00E-07** |
| N,N-Dimethylglycine | 0,006 (0,002) | 0,007 (0,002) | 0,006 (0,003) | 0,006 (0,003) | 0,002 (0,001) | 2,49E-01 | 8,75E-01 | 3,30E-01 | **5,73E-10** |
| Phenylalanine | 0,054 (0,019) | 0,055 (0,015) | 0,052 (0,015) | 0,054 (0,016) | 0,110 (0,025) | 8,79E-01 | 7,23E-01 | 1,00E+00 | **5,69E-11** |
| Tyrosine | 0,049 (0,015) | 0,058 (0,024) | 0,057 (0,016) | 0,055 (0,02) | 0,059 (0,016) | 1,27E-01 | 1,20E-01 | 2,06E-01 | **1,36E-03** |
| Valine | 0,225 (0,059) | 0,242 (0,054) | 0,25 (0,053) | 0,245 (0,067) | 0,258 (0,032) | 3,47E-01 | 9,48E-02 | 3,04E-01 | **3,68E-03** |
| Acetic acid | 0,023 (0,011) | 0,024 (0,015) | 0,017 (0,009) | 0,018 (0,01) | 0,077 (0,035) | 9,46E-01 | **1,97E-02** | **2,78E-02** | **1,01E-10** |
| Citric acid | 0,168 (0,066) | 0,148 (0,063) | 0,149 (0,068) | 0,162 (0,055) | 0,058 (0,071) | 1,50E-01 | 2,11E-01 | 3,83E-01 | **2,86E-07** |
| Formic acid | 0,020 (0,01) | 0,022 (0,011) | 0,019 (0,012) | 0,019 (0,009) | 0,048 (0,017) | 8,14E-01 | 5,59E-01 | 4,50E-01 | **2,97E-09** |
| Lactic acid | 2,103 (1,306) | 1,924 (0,977) | 2,394 (0,932) | 2,334 (0,959) | 3,086 (1,255) | 7,73E-01 | 7,85E-02 | 1,02E-01 | **1,13E-04** |
| Succinic acid | 0,007 (0,006) | 0,004 (0,003) | 0,005 (0,006) | 0,006 (0,009) | 0,013 (0,007) | 7,83E-02 | 1,68E-01 | 1,71E-01 | **5,68E-05** |
| BHB | 0,232 (0,24) | 0,122 (0,148) | 0,100 (0,121) | 0,125 (0,197) | 0,117 (0,126) | **4,36E-03** | **5,40E-04** | **1,27E-03** | **3,08E-03** |
| Acetoacetate | 0,031 (0,032) | 0,013 (0,01) | 0,01 (0,012) | 0,031 (0,095) | 0,007 (0,004) | 6,28E-02 | **3,88E-03** | 8,74E-02 | **3,66E-04** |
| Acetone | 0,041 (0,024) | 0,027 (0,014) | 0,023 (0,014) | 0,032 (0,031) | 0,051 (0,043) | **9,06E-03** | **5,68E-04** | **1,62E-02** | 6,50E-01 |
| Pyruvate | 0,08 (0,041) | 0,087 (0,032) | 0,104 (0,051) | 0,115 (0,046) | 0,022 (0,012) | 1,92E-01 | **4,06E-02** | **8,55E-04** | **5,65E-11** |
| Glucose | 5,246 (1,021) | 5,346 (1,037) | 5,465 (0,84) | 5,576 (1,646) | 5,493 (1,175) | 7,33E-01 | 2,01E-01 | 2,84E-01 | 2,21E-01 |
| Dimethylsulfone | 0,013 (0,007) | 0,061 (0,258) | 0,107 (0,343) | 0,012 (0,006) | 0,013 (0,007) | 9,76E-01 | 1,33E-01 | 6,95E-01 | 5,17E-01 |
| TPTG | 113,645 (57,52) | 100,557 (36,784) | 99,015 (43,533) | 99,262 (45,611) | 96,967 (39,684) | 4,62E-01 | 4,64E-01 | 1,58E-01 | 2,08E-01 |
| TPCH | 204,351 (49,204) | 228,318 (46,909) | 214,806 (40,405) | 204,636 (50,525) | 229,306 (53,879) | **4,30E-02** | 2,58E-01 | 9,00E-01 | 5,49E-02 |
| LDCH | 105,406 (34,872) | 125,7 (32,447) | 120,366 (33,832) | 115,226 (37,575) | 138,903 (41,349) | **2,16E-02** | 1,05E-01 | 3,79E-01 | **1,51E-03** |
| HDCH | 63,625 (19,918) | 70,786 (19,034) | 65,061 (14,661) | 61,895 (16,689) | 57,297 (12,816) | 8,26E-02 | 4,83E-01 | 7,73E-01 | 2,48E-01 |
| TPA1 | 160,537 (32,583) | 171,859 (28,326) | 161,201 (24,527) | 151,098 (28,44) | 146,924 (23,316) | 1,03E-01 | 5,79E-01 | 2,90E-01 | 1,18E-01 |
| TPA2 | 33,911 (7,572) | 37,965 (6,269) | 34,933 (5,138) | 33,002 (4,887) | 33,637 (5,482) | **2,16E-02** | 4,64E-01 | 7,84E-01 | 9,01E-01 |
| TPAB | 82,001 (19,898) | 89,706 (19,48) | 85,423 (20,173) | 84,048 (22,918) | 100,939 (25,706) | 2,72E-01 | 7,39E-01 | 7,96E-01 | **6,30E-03** |
| LDHD | 1,706 (0,465) | 1,843 (0,516) | 1,914 (0,62) | 1,893 (0,507) | 2,478 (0,73) | 5,00E-01 | 2,09E-01 | 3,29E-01 | **1,55E-05** |
| ABA1 | 0,52 (0,132) | 0,532 (0,132) | 0,54 (0,145) | 0,566 (0,155) | 0,692 (0,163) | 8,26E-01 | 5,25E-01 | 4,29E-01 | **1,12E-04** |
| TBPN | 1,491 (0,362) | 1,631 (0,354) | 1,553 (0,367) | 1,528 (0,417) | 1,835 (0,467) | 2,72E-01 | 7,39E-01 | 7,96E-01 | **6,30E-03** |
| VLPN | 0,145 (0,082) | 0,127 (0,06) | 0,124 (0,059) | 0,129 (0,054) | 0,121 (0,063) | 3,36E-01 | 4,12E-01 | 4,04E-01 | 1,62E-01 |
| IDPN | 0,087 (0,038) | 0,091 (0,038) | 0,082 (0,03) | 0,075 (0,028) | 0,121 (0,046) | 8,02E-01 | 6,31E-01 | 1,37E-01 | **7,75E-03** |
| LDPN | 1,215 (0,316) | 1,386 (0,319) | 1,325 (0,336) | 1,31 (0,384) | 1,492 (0,399) | 5,32E-02 | 2,58E-01 | 4,12E-01 | **8,75E-03** |
| L1PN | 0,257 (0,073) | 0,272 (0,077) | 0,248 (0,063) | 0,245 (0,088) | 0,28 (0,097) | 3,43E-01 | 6,63E-01 | 3,04E-01 | 3,82E-01 |
| L2PN | 0,172 (0,084) | 0,206 (0,098) | 0,198 (0,075) | 0,203 (0,095) | 0,282 (0,101) | 1,80E-01 | 2,46E-01 | 2,84E-01 | **8,53E-05** |
| L3PN | 0,173 (0,073) | 0,205 (0,064) | 0,199 (0,082) | 0,197 (0,09) | 0,283 (0,094) | **4,14E-02** | 2,28E-01 | 3,40E-01 | **1,17E-05** |
| L4PN | 0,148 (0,068) | 0,186 (0,093) | 0,176 (0,095) | 0,165 (0,088) | 0,214 (0,091) | 1,22E-01 | 4,46E-01 | 6,95E-01 | **5,55E-03** |
| L5PN | 0,163 (0,079) | 0,203 (0,105) | 0,199 (0,085) | 0,183 (0,076) | 0,22 (0,071) | 1,11E-01 | 1,37E-01 | 3,87E-01 | **5,55E-03** |

| | | | | | | | | | |
|---|---|---|---|---|---|---|---|---|---|
| **L6PN** | 0,303 (0,114) | 0,313 (0,110) | 0,31 (0,095) | 0,303 (0,111) | 0,278 (0,099) | 5,00E-01 | 3,63E-01 | 9,59E-01 | 5,49E-01 |
| **VLTG** | 71,304 (46,955) | 62,284 (30,014) | 63,232 (34,993) | 62,19 (36,447) | 56,198 (27,27) | 5,29E-01 | 7,28E-01 | 3,48E-01 | 1,98E-01 |
| **IDTG** | 10,25 (8,619) | 8,71 (5,432) | 8,703 (6,905) | 8,456 (7,532) | 5,059 (5,368) | 6,44E-01 | 6,00E-01 | 2,94E-01 | **2,69E-03** |
| **LDTG** | 19,093 (3,649) | 18,829 (4,676) | 18,165 (4,273) | 18,535 (7,418) | 22,602 (5,435) | 4,30E-01 | 1,74E-01 | 1,15E-01 | **1,20E-02** |
| **HDTG** | 11,218 (2,904) | 10,454 (2,998) | 10,121 (3,488) | 9,186 (2,616) | 11,647 (3,234) | 1,35E-01 | 1,43E-01 | **7,13E-03** | 6,70E-01 |
| **VLCH** | 19,345 (12,546) | 17,193 (9,892) | 16,762 (9,176) | 16,04 (7,736) | 18,385 (10,478) | 6,55E-01 | 6,00E-01 | 4,16E-01 | 8,31E-01 |
| **IDCH** | 11,722 (6,407) | 12,222 (6,272) | 11,100 (5,016) | 9,51 (4,268) | 18,09 (8,085) | 8,62E-01 | 7,79E-01 | 1,37E-01 | **3,38E-03** |
| **VLFC** | 8,661 (4,973) | 7,489 (3,699) | 7,321 (3,789) | 7,091 (3,605) | 7,508 (3,887) | 3,75E-01 | 3,48E-01 | 1,83E-01 | 3,46E-01 |
| **IDFC** | 3,556 (1,949) | 3,719 (1,825) | 3,309 (1,513) | 2,900 (1,288) | 4,714 (2,289) | 9,34E-01 | 6,90E-01 | 1,19E-01 | 6,82E-02 |
| **LDFC** | 31,636 (9,532) | 36,14 (8,494) | 34,596 (9,178) | 33,089 (10,301) | 44,132 (11,279) | **4,45E-02** | 1,81E-01 | 6,31E-01 | **5,06E-05** |
| **HDFC** | 13,512 (5,338) | 14,713 (4,401) | 13,382 (4,292) | 12,606 (4,471) | 17,08 (3,471) | 3,21E-01 | 9,12E-01 | 4,60E-01 | **1,51E-03** |
| **VLPL** | 19,188 (11,102) | 16,873 (8,158) | 16,968 (8,681) | 16,517 (7,83) | 13,97 (7,408) | 4,62E-01 | 6,31E-01 | 3,40E-01 | **4,52E-02** |
| **IDPL** | 7,596 (3,901) | 7,987 (3,062) | 7,606 (2,999) | 6,867 (3,076) | 6,682 (3,194) | 7,33E-01 | 8,13E-01 | 4,33E-01 | 4,13E-01 |
| **LDPL** | 61,14 (16,726) | 70,324 (15,084) | 67,079 (16,039) | 65,249 (18,464) | 74,862 (19,635) | **2,34E-02** | 1,02E-01 | 3,95E-01 | **4,99E-03** |
| **HDPL** | 87,658 (22,645) | 93,17 (21,791) | 84,705 (18,055) | 81,865 (18,94) | 73,239 (15,501) | 2,55E-01 | 7,17E-01 | 3,04E-01 | **1,32E-02** |
| **HDA1** | 159,966 (35,214) | 171,878 (31,708) | 160,355 (26,061) | 150,399 (30,56) | 147,738 (24,729) | 1,17E-01 | 7,39E-01 | 2,71E-01 | 2,34E-01 |
| **HDA2** | 34,005 (6,967) | 37,945 (6,095) | 34,974 (4,739) | 33,326 (4,561) | 33,502 (5,245) | **2,12E-02** | 4,08E-01 | 8,53E-01 | 8,10E-01 |
| **VLAB** | 7,987 (4,49) | 6,976 (3,287) | 6,844 (3,268) | 7,096 (2,987) | 6,658 (3,441) | 3,32E-01 | 4,08E-01 | 4,04E-01 | 1,67E-01 |
| **IDAB** | 4,793 (2,09) | 5,029 (2,105) | 4,519 (1,677) | 4,139 (1,554) | 6,665 (2,548) | 8,08E-01 | 6,26E-01 | 1,37E-01 | **7,90E-03** |
| **LDAB** | 66,832 (17,398) | 76,211 (17,544) | 72,852 (18,457) | 72,03 (21,146) | 82,033 (21,922) | 5,32E-02 | 2,58E-01 | 4,12E-01 | **8,75E-03** |
| **V1TG** | 31,179 (28,166) | 26,287 (15,853) | 26,922 (21,549) | 27,771 (25,498) | 30,007 (14,355) | 6,88E-01 | 6,00E-01 | 4,73E-01 | 7,26E-01 |
| **V2TG** | 12,09 (8,693) | 10,392 (6,135) | 11,303 (6,811) | 11,23 (6,396) | 8,522 (4,779) | 6,60E-01 | 9,88E-01 | 8,65E-01 | 1,12E-01 |
| **V3TG** | 10,948 (7,371) | 9,624 (5,923) | 9,87 (5,443) | 9,515 (4,755) | 9,14 (5,025) | 5,39E-01 | 9,06E-01 | 6,20E-01 | 4,70E-01 |
| **V4TG** | 8,894 (4,561) | 8,018 (4,139) | 7,564 (3,678) | 7,326 (2,572) | 7,114 (3,813) | 3,51E-01 | 2,14E-01 | 2,52E-01 | 7,47E-02 |
| **V5TG** | 2,519 (0,77) | 2,373 (0,883) | 2,039 (0,759) | 2,265 (0,646) | 2,373 (0,813) | 1,24E-01 | **8,68E-03** | 9,62E-02 | 2,34E-01 |
| **V1CH** | 6,193 (5,237) | 4,821 (3,25) | 4,988 (3,969) | 4,894 (4,093) | 5,542 (3,345) | 3,79E-01 | 4,60E-01 | 2,09E-01 | 9,78E-01 |
| **V2CH** | 2,893 (2,154) | 2,606 (1,656) | 2,706 (1,631) | 2,521 (1,413) | 2,288 (1,517) | 8,26E-01 | 9,59E-01 | 9,53E-01 | 4,33E-01 |
| **V3CH** | 3,789 (2,767) | 3,251 (2,391) | 3,181 (1,966) | 2,839 (1,701) | 3,831 (2,184) | 4,81E-01 | 5,39E-01 | 2,14E-01 | 7,57E-01 |
| **V4CH** | 4,981 (2,791) | 4,741 (2,909) | 4,248 (2,314) | 3,813 (1,514) | 5,232 (2,784) | 6,71E-01 | 3,71E-01 | 1,21E-01 | 7,67E-01 |
| **V5CH** | 1,37 (0,613) | 1,294 (0,679) | 0,992 (0,565) | 1,199 (0,544) | 0,575 (0,586) | 4,90E-01 | **1,38E-02** | 2,49E-01 | **8,06E-06** |
| **V1FC** | 2,558 (2,287) | 2,092 (1,31) | 2,269 (1,833) | 2,14 (1,981) | 0,934 (1,006) | 5,75E-01 | 8,77E-01 | 3,55E-01 | **1,09E-03** |
| **V2FC** | 1,263 (1,092) | 1,059 (0,769) | 1,149 (0,781) | 1,124 (0,729) | 1,104 (0,792) | 7,22E-01 | 9,65E-01 | 1,00E+00 | 8,42E-01 |
| **V3FC** | 1,652 (1,292) | 1,344 (0,981) | 1,338 (0,87) | 1,284 (0,843) | 1,389 (0,907) | 4,53E-01 | 5,01E-01 | 2,74E-01 | 5,13E-01 |
| **V4FC** | 2,119 (1,396) | 1,980 (1,41) | 1,716 (1,06) | 1,546 (0,782) | 2,559 (1,488) | 6,22E-01 | 2,87E-01 | 8,23E-02 | 2,77E-01 |
| **V5FC** | 0,487 (0,328) | 0,303 (0,353) | 0,22 (0,304) | 0,226 (0,269) | 0,398 (0,407) | **2,01E-02** | **1,11E-03** | **3,25E-03** | 1,45E-01 |
| **V1PL** | 5,766 (4,72) | 4,858 (2,746) | 5,122 (3,71) | 5,242 (3,973) | 4,555 (2,266) | 6,38E-01 | 6,36E-01 | 6,10E-01 | 4,25E-01 |
| **V2PL** | 3,292 (2,267) | 2,863 (1,604) | 3,143 (1,771) | 3,06 (1,605) | 2,163 (1,200) | 6,44E-01 | 9,35E-01 | 8,24E-01 | **4,99E-02** |
| **V3PL** | 3,917 (2,703) | 3,554 (2,038) | 3,62 (1,919) | 3,413 (1,597) | 2,93 (1,707) | 6,93E-01 | 9,76E-01 | 5,49E-01 | 1,73E-01 |
| **V4PL** | 4,435 (2,284) | 4,169 (2,181) | 3,88 (1,808) | 3,677 (1,251) | 4,355 (2,184) | 5,70E-01 | 3,67E-01 | 2,09E-01 | 7,62E-01 |
| **V5PL** | 1,581 (0,768) | 1,469 (0,758) | 1,197 (0,65) | 1,377 (0,63) | 1,017 (0,722) | 3,55E-01 | 6,14E-02 | 3,11E-01 | **3,94E-03** |
| **L1TG** | 6,515 (1,808) | 5,963 (2,059) | 5,411 (1,768) | 5,844 (2,859) | 6,051 (2,381) | 2,22E-01 | **1,84E-02** | **4,51E-02** | 3,22E-01 |
| **L2TG** | 2,576 (0,732) | 2,781 (0,803) | 2,619 (0,681) | 2,69 (1,279) | 2,957 (0,915) | 2,03E-01 | 7,45E-01 | 3,67E-01 | 8,66E-02 |
| **L3TG** | 2,067 (0,654) | 2,193 (0,738) | 2,032 (0,533) | 2,16 (0,797) | 2,883 (0,622) | 3,32E-01 | 8,48E-01 | 9,06E-01 | **1,45E-05** |
| **L4TG** | 1,944 (0,82) | 2,152 (1,234) | 2,042 (0,87) | 2,071 (1,309) | 2,528 (1,112) | 5,00E-01 | 7,34E-01 | 9,47E-01 | 5,06E-02 |
| **L5TG** | 2,092 (0,905) | 2,234 (1,261) | 2,23 (0,965) | 2,167 (1,157) | 2,07 (0,931) | 7,39E-01 | 5,95E-01 | 9,29E-01 | 8,53E-01 |
| **L6TG** | 3,546 (1,044) | 3,337 (0,929) | 3,348 (1,024) | 3,551 (1,273) | 4,138 (1,275) | 4,44E-01 | 6,26E-01 | 7,67E-01 | **4,03E-02** |
| **L1CH** | 26,664 (8,601) | 29,307 (8,905) | 26,852 (7,082) | 25,793 (9,262) | 28,878 (11,509) | 2,22E-01 | 8,59E-01 | 6,00E-01 | 4,87E-01 |
| **L2CH** | 17,133 (9,663) | 21,037 (11,35) | 20,367 (8,518) | 20,454 (10,598) | 28,331 (10,844) | 1,39E-01 | 1,76E-01 | 2,64E-01 | **9,56E-05** |
| **L3CH** | 15,891 (8,19) | 19,747 (7,257) | 19,159 (8,916) | 18,64 (9,469) | 26,54 (9,674) | **3,13E-02** | 1,30E-01 | 3,18E-01 | **3,99E-05** |
| **L4CH** | 12,732 (6,167) | 16,676 (7,811) | 15,83 (9,012) | 14,627 (8,443) | 18,57 (8,223) | 5,51E-02 | 3,95E-01 | 5,20E-01 | **4,68E-03** |

| | | | | | | | | | |
|---|---|---|---|---|---|---|---|---|---|
| **L5CH** | 12,217 (6,347) | 15,995 (8,442) | 15,67 (7,182) | 14,255 (6,52) | 16,844 (5,793) | 9,24E-02 | 7,85E-02 | 2,94E-01 | **6,57E-03** |
| **L6CH** | 20,449 (7,93) | 22,000 (7,699) | 21,934 (6,647) | 21,155 (7,348) | 18,863 (7,104) | 2,82E-01 | 2,17E-01 | 6,05E-01 | 5,31E-01 |
| **L1FC** | 8,334 (2,624) | 9,044 (2,562) | 8,297 (1,97) | 7,911 (2,934) | 9,232 (3,296) | 2,34E-01 | 8,02E-01 | 4,08E-01 | 3,35E-01 |
| **L2FC** | 5,828 (3,14) | 7,085 (3,389) | 6,644 (2,588) | 6,619 (3,455) | 9,671 (3,196) | 9,53E-02 | 2,14E-01 | 4,33E-01 | **1,87E-05** |
| **L3FC** | 5,589 (2,631) | 6,627 (2,221) | 6,285 (2,387) | 6,068 (2,812) | 8,185 (2,536) | 7,48E-02 | 2,06E-01 | 6,20E-01 | **2,80E-04** |
| **L4FC** | 4,398 (1,772) | 5,428 (1,729) | 5,111 (2,157) | 4,801 (2,13) | 6,497 (2,062) | **3,92E-02** | 3,01E-01 | 7,28E-01 | **1,27E-04** |
| **L5FC** | 3,855 (1,506) | 4,672 (2,037) | 4,529 (1,625) | 4,119 (1,485) | 5,359 (1,475) | 1,10E-01 | 1,58E-01 | 7,39E-01 | **3,74E-04** |
| **L6FC** | 5,609 (2,151) | 6,024 (1,95) | 5,942 (1,662) | 5,603 (1,806) | 5,735 (2,056) | 2,40E-01 | 2,71E-01 | 7,12E-01 | 6,35E-01 |
| **L1PL** | 15,272 (4,309) | 16,369 (4,453) | 15,024 (3,507) | 14,638 (4,834) | 16,161 (5,738) | 3,03E-01 | 8,82E-01 | 4,29E-01 | 6,25E-01 |
| **L2PL** | 9,839 (4,922) | 11,788 (5,577) | 11,328 (4,251) | 11,451 (5,388) | 15,375 (5,387) | 1,20E-01 | 2,06E-01 | 3,59E-01 | **1,10E-04** |
| **L3PL** | 9,152 (4,064) | 11,008 (3,457) | 10,607 (4,307) | 10,434 (4,705) | 14,374 (4,765) | **3,57E-02** | 1,60E-01 | 3,75E-01 | **4,24E-05** |
| **L4PL** | 7,405 (3,249) | 9,334 (4,06) | 8,863 (4,472) | 8,299 (4,132) | 10,245 (4,114) | 6,77E-02 | 4,12E-01 | 5,49E-01 | **1,16E-02** |
| **L5PL** | 6,929 (3,182) | 8,804 (4,228) | 8,625 (3,572) | 8,003 (3,135) | 9,21 (2,879) | 5,51E-02 | 8,91E-02 | 2,74E-01 | **7,28E-03** |
| **L6PL** | 11,968 (3,898) | 12,652 (3,688) | 12,37 (3,235) | 12,116 (3,613) | 10,668 (3,707) | 3,28E-01 | 4,04E-01 | 7,51E-01 | 2,26E-01 |
| **L1AB** | 14,143 (4,018) | 14,984 (4,219) | 13,646 (3,444) | 13,48 (4,853) | 15,414 (5,325) | 3,47E-01 | 6,57E-01 | 3,08E-01 | 3,78E-01 |
| **L2AB** | 9,47 (4,637) | 11,312 (5,384) | 10,885 (4,112) | 11,146 (5,228) | 15,493 (5,549) | 1,77E-01 | 2,43E-01 | 2,84E-01 | **8,53E-05** |
| **L3AB** | 9,499 (4,016) | 11,289 (3,542) | 10,954 (4,52) | 10,853 (4,953) | 15,541 (5,17) | **4,14E-02** | 2,31E-01 | 3,40E-01 | **1,17E-05** |
| **L4AB** | 8,155 (3,764) | 10,249 (5,121) | 9,658 (5,206) | 9,048 (4,86) | 11,774 (4,979) | 1,22E-01 | 4,46E-01 | 6,95E-01 | **5,55E-03** |
| **L5AB** | 8,944 (4,361) | 11,169 (5,752) | 10,924 (4,649) | 10,059 (4,153) | 12,09 (3,931) | 1,11E-01 | 1,37E-01 | 3,87E-01 | **5,55E-03** |
| **L6AB** | 16,684 (6,284) | 17,21 (6,068) | 17,064 (5,201) | 16,691 (6,083) | 15,293 (5,423) | 5,00E-01 | 3,63E-01 | 9,59E-01 | 5,49E-01 |
| **H1TG** | 3,984 (1,196) | 3,617 (1,815) | 3,58 (1,568) | 3,077 (1,484) | 4,268 (1,197) | **4,70E-02** | 1,41E-01 | **7,44E-03** | 2,86E-01 |
| **H2TG** | 1,892 (0,619) | 1,776 (0,651) | 1,695 (0,685) | 1,563 (0,626) | 2,287 (0,652) | 1,82E-01 | 2,37E-01 | **2,28E-02** | **2,86E-02** |
| **H3TG** | 2,18 (0,766) | 2,088 (0,711) | 1,956 (0,789) | 1,889 (0,621) | 2,106 (0,688) | 2,52E-01 | 1,96E-01 | **3,99E-02** | 5,13E-01 |
| **H4TG** | 3,471 (1,179) | 3,311 (1,124) | 3,191 (1,149) | 3,037 (0,921) | 2,969 (1,138) | 5,19E-01 | 2,20E-01 | **4,84E-02** | 6,93E-02 |
| **H1CH** | 21,482 (10,826) | 23,076 (11,737) | 22,014 (8,771) | 20,466 (9,899) | 19,664 (7,897) | 7,16E-01 | 5,01E-01 | 7,17E-01 | 8,10E-01 |
| **H2CH** | 9,559 (3,55) | 10,816 (4,065) | 9,637 (2,941) | 9,166 (3,228) | 9,002 (2,486) | 1,85E-01 | 6,00E-01 | 7,84E-01 | 7,46E-01 |
| **H3CH** | 11,468 (3,165) | 12,809 (2,682) | 11,286 (2,38) | 11,012 (2,598) | 9,735 (2,15) | 6,11E-02 | 9,23E-01 | 5,49E-01 | **2,23E-02** |
| **H4CH** | 21,002 (5,6) | 23,343 (4,435) | 21,631 (3,799) | 20,899 (5,062) | 17,856 (3,664) | 8,26E-02 | 6,68E-01 | 9,12E-01 | **1,07E-02** |
| **H1FC** | 4,851 (2,88) | 5,541 (2,628) | 5,183 (2,374) | 4,608 (2,387) | 6,546 (2,056) | 2,43E-01 | 3,99E-01 | 8,02E-01 | **2,25E-03** |
| **H2FC** | 2,171 (0,968) | 2,497 (0,866) | 2,169 (0,831) | 2,01 (0,778) | 2,82 (0,632) | 1,65E-01 | 8,77E-01 | 5,79E-01 | **1,96E-03** |
| **H3FC** | 2,23 (0,978) | 2,681 (0,74) | 2,336 (0,701) | 2,093 (0,763) | 2,734 (0,704) | **4,62E-02** | 6,26E-01 | 4,55E-01 | **1,76E-02** |
| **H4FC** | 3,971 (1,457) | 4,339 (1,057) | 3,888 (1,054) | 3,585 (1,196) | 4,229 (1,04) | 3,47E-01 | 4,12E-01 | 1,93E-01 | 8,10E-01 |
| **H1PL** | 25,372 (12,591) | 26,395 (13,824) | 24,979 (10,712) | 23,14 (11,238) | 23,712 (8,954) | 8,97E-01 | 9,35E-01 | 4,12E-01 | 9,73E-01 |
| **H2PL** | 14,53 (4,728) | 15,826 (5,562) | 13,977 (4,084) | 13,58 (4,438) | 13,119 (3,413) | 4,00E-01 | 8,48E-01 | 5,06E-01 | 2,99E-01 |
| **H3PL** | 18,157 (4,189) | 19,669 (4,172) | 17,17 (3,414) | 17,065 (3,881) | 13,481 (3,106) | 1,35E-01 | 6,41E-01 | 3,37E-01 | **1,21E-05** |
| **H4PL** | 29,037 (5,507) | 30,898 (4,775) | 28,327 (3,933) | 27,923 (5,37) | 22,047 (3,987) | 1,87E-01 | 6,10E-01 | 5,84E-01 | **1,32E-06** |
| **H1A1** | 30,38 (17,575) | 32,337 (20,7) | 31,125 (15,3) | 27,357 (16,037) | 30,768 (12,405) | 9,34E-01 | 7,73E-01 | 3,95E-01 | 3,86E-01 |
| **H2A1** | 19,691 (5,647) | 21,283 (5,625) | 19,248 (4,75) | 18,485 (4,54) | 17,656 (3,968) | 2,92E-01 | 9,59E-01 | 4,51E-01 | 2,29E-01 |
| **H3A1** | 28,802 (6,397) | 31,214 (5,949) | 27,507 (5,004) | 26,963 (5,754) | 25,442 (5,225) | 1,17E-01 | 6,20E-01 | 3,63E-01 | 5,15E-02 |
| **H4A1** | 78,516 (16,635) | 84,086 (13,303) | 79,369 (11,273) | 75,522 (15,132) | 70,962 (12,164) | 1,61E-01 | 8,30E-01 | 6,41E-01 | 7,03E-02 |
| **H1A2** | 2,684 (1,693) | 2,984 (1,866) | 2,733 (1,508) | 2,458 (1,448) | 2,982 (1,215) | 6,17E-01 | 7,56E-01 | 6,73E-01 | 2,10E-01 |
| **H2A2** | 3,609 (1,191) | 4,059 (1,398) | 3,627 (1,004) | 3,407 (1,056) | 3,326 (0,859) | 2,00E-01 | 9,47E-01 | 5,39E-01 | 4,83E-01 |
| **H3A2** | 6,744 (1,625) | 7,719 (1,779) | 6,869 (1,324) | 6,666 (1,284) | 6,130 (1,190) | **3,07E-02** | 6,41E-01 | 8,07E-01 | 1,04E-01 |
| **H4A2** | 19,778 (5,062) | 21,991 (4,427) | 20,514 (3,467) | 19,524 (4,28) | 18,526 (3,688) | 5,91E-02 | 5,49E-01 | 9,65E-01 | 2,37E-01 |

The metabolites are expressed in mmol/L. Lipoprotein values from TPTG to TPAB are expressed in mg/dL. The other lipoprotein values are expressed in nmol/L, except for the LDHD and ABA1 values, which are not expressed with a unit (ratio). Significant p values of less than 0.05 are indicated in bold.

Supplemental Table S4

**Table S4: Evaluation of the performances of different combinations of the IVDr-6M-BB with the addition of the three compounds citrate, HDTG, and V5FC.**

| | | | Train data (PD vs HC) | | | | | Prediction of data (HC, PD, MSA, PSP) | | | | |
|---|---|---|---|---|---|---|---|---|---|---|---|---|
| **Models** | **AUC (95%CI)** | **Threshold** | **Sensitivity (%)** | **Specificity (%)** | **PPV (%)** | **NPV (%)** | **Accuracy (%)** | **Sensitivity (%)** | **Specificity (%)** | **PPV (%)** | **NPV (%)** | **Accuracy (%)** |
| IVDr-6M-BB + Citrate + V5FC | 0.959 (0.91-1) | 0.494 | 96.7 | 93.1 | 93.5 | 96.4 | 94.9 | 96.7 | 80.9 | 63.0 | 98.6 | 84.9 |
| IVDr-6M-BB + Citrate + HDTG + V5FC | 0.990 (0.97-1) | 0.445 | 96.7 | 93.1 | 93.5 | 96.4 | 94.9 | 96.7 | 76.4 | 58.0 | 98.6 | 81.5 |
| IVDr-6M-BB + V5FC | 0.914 (0.84-0.99) | 0.646 | 80.0 | 96.6 | 96.0 | 82.4 | 88.1 | 80.0 | 82.0 | 60.0 | 92.4 | 81.5 |
| IVDr-6M-BB + Citrate + HDTG | 0.984 (0.96-1) | 0.377 | 93.3 | 93.1 | 93.3 | 93.1 | 93.2 | 93.3 | 75.3 | 56.0 | 97.1 | 79.8 |
| IVDr-6M-BB + Citrate | 0.940 (0.88-1) | 0.471 | 90.0 | 86.2 | 87.1 | 89.3 | 88.1 | 90.0 | 73.0 | 52.9 | 95.6 | 77.3 |
| IVDr-6M-BB + HDTG + V5FC | 0.938 (0.87-1) | 0.435 | 90.0 | 89.7 | 90.0 | 89.7 | 89.8 | 90.0 | 73.0 | 52.9 | 95.6 | 77.3 |
| IVDr-6M-BB + HDTG | 0.928 (0.86-1) | 0.428 | 90.0 | 89.7 | 90.0 | 89.7 | 89.8 | 90.0 | 71.9 | 51.9 | 95.5 | 76.5 |
| IVDr-6M-BB | 0.878 (0.78-0.97) | 0.510 | 83.3 | 86.2 | 86.2 | 83.3 | 84.7 | 83.3 | 74.2 | 52.0 | 93.0 | 76.5 |

ROC curve analyses of the different logistic regression models based on PD (n=30), HC (n=29), MSA (n=30) and PSP (n=30) each including the 6 core metabolites (acetoacetate, BHB, betaine, creatine, pyruvate, valine) plus additional variables. Models are listed in descending order of accuracy of the data set including all cases. *AUC: Area Under the Curve. CI: Confident Interval. PPV: positive predictive value; NPV: negative predictive value.*

Supplemental Table S5

**Table S5: List of metabolites identified in serum samples at 950 MHz**

| | Metabolite | Group | δ (ppm) | Multiplicity |
|---|---|---|---|---|
| **1** | **3-hydroxybutyrate** | γ-$CH_3$ | 1.19 | d |
| | | half α-$CH_2$ | 2.30 | dd |
| | | half α-$CH_2$ | 2.39 | dd |
| | | β-CH | 4.15 | m |
| **2** | **Acetoacetate** | | 3.44 | s |
| | | | 2.27 | s |
| **3** | **Acetate** | $CH_3$ | 1.91 | s |
| **4** | **Acetone** | $CH_2CO$ | 2.22 | s |
| **5** | **Alanine** | $CH_3$ | 1.47 | d |
| | | α-CH | 3.77 | q |
| **6** | **Albumin lysyl** | ϵ-$CH_2$ | 2.88 | t |
| | | ϵ-$CH_2$ | 2.95 | t |
| | | ϵ-$CH_2$ | 3.01 | t |
| **7** | **Arginine** | γ-$CH_2$ | 1.64 | |
| | | γ-$CH_2$ | 1.69 | |
| | | β-$CH_2$ | 1.89 | |
| | | δ-$CH_2$ | 3.23 | t |
| **8** | **Aspartate** | half β-$CH_2$ | 2.66 | q |
| | | half β-$CH_2$ | 2.80 | dd |
| **9** | **Betaine** | $CH_3$ | 3.26 | s |
| | | $CH_2$ | 3.90 | |
| **10** | **Choline** | N $(CH_3)_3$ | 3.21 | s |
| | | | 3.51 | |
| | | | 4.06 | |
| **11** | **Citrate** | half $CH_2$ | 2.53 | d |
| | | half $CH_2$ | 2.66 | d |
| **12** | **Creatine** | $CH_3$ | 3.03 | s |
| | | $CH_2$ | 3.92 | s |
| **13** | **Creatinine** | $CH_3$ | 3.04 | s |
| | | $CH_2$ | 4.05 | s |
| **14** | **Dimethylamine** | $CH_3$ | 2.71 | s |
| **15** | **Ethanol** | $CH_3$ | 1.17 | t |
| | | $CH_3COH$ | 3.65 | q |
| **16** | **Fatty acids** | $\mathit{CH_3}CH_2$ | 0.93 | m |
| | | $CH_3\mathit{CH_2}(CH_2)n$ | 1.24 | m |
| | | $CH_3\mathit{CH_2}(CH_2)n$ | 1.26 | m |
| | | $CH_2$ | 1.26 | m |
| | | $CH_2$ | 1.30 | m |
| | | $\mathit{CH_2}CH_2C=C$ | 1.68 | |
| | | $CH_2C=C$ | 2.00 | m |
| | | $CH_2CO$ | 2.22 | m |
| | | $C=CCH_2C=C$ | 2.72 | m |
| | | CH=*CH*CH2*CH*=CH | 5.26 | m |
| | | CH=*CH*CH2*CH*=CH | 5.29 | m |
| **17** | **Formate** | CH | 8.45 | s |
| **18** | **Fructose** | | 3.99 | m |
| | | | 4.01 | dd |
| **19** | **Fucose / β-Galactose** | | 4.54 | d |
| **20** | **Glucose** | H2 | 3.24 | t |
| | | H4 | 3.40 | t |
| | | H4 | 3.41 | t |
| | | H5 | 3.46 | m |
| | | H3 | 3.48 | t |
| | | H2 | 3.53 | q |
| | | H3 | 3.71 | t |
| | | half $CH_2$-C6 | 3.72 | q |
| | | half $CH_2$-C6 | 3.76 | m |
| | | H5 | 3.82 | dd |
| | | half $CH_2$-C6 | 3.84 | m |
| | | half $CH_2$-C6 | 3.89 | dd |
| | | H1 | 4.64 | d |
| | | H1 | 5.23 | d |
| **21** | **Glutamate** | half β-$CH_2$ | 2.04 | m |
| | | half β-$CH_2$ | 2.12 | m |
| | | half γ-$CH_2$ | 2.34 | m |
| | | half γ-$CH_2$ | 2.36 | m |
| | | | 3.74 | m |
| **22** | **Glutamine** | | 2.08 | |
| | | | 2.09 | |
| | | half β-$CH_2$ | 2.11 | m |
| | | half γ-$CH_2$ | 2.44 | m |
| | | | 2.46 | m |
| | | | 3.74 | |
| **23** | **Glycerol** | half $CH_2$ | 3.56 | q |
| | | half $CH_2$ | 3.65 | q |
| | | $C_2$-H | 3.87 | m |
| **24** | **Glycerol backbone** | | 4.06 | |
| **25** | **Glycerophosphocholine** | | 3.22 | s |
| | | $NCH_2$ | 3.66 | m |

| | | | | |
|---|---|---|---|---|
| | | $OCH_2$ | 4.29 | m |
| **26** | **Glycine** | $CH_2$ | 3.55 | s |
| **27** | **Histidine** | | 3.09 | dd |
| | | | 3.98 | dd |
| | | H4 | 7.04 | s |
| | | H2 | 7.75 | s |
| **28** | **Isoleucine** | δ-$CH_3$ | 0.93 | t |
| | | β-CH3 | 1.00 | d |
| | | half ɣ-$CH_2$ | 1.24 | |
| | | half ɣ-$CH_2$ | 1.46 | |
| | | | 1.96 | |
| | | | 3.65 | |
| **29** | **Lactate** | $CH_3$ | 1.32 | d |
| | | CH | 4.11 | q |
| **30** | **Lactose** | | 3.55 | |
| | | | 3.66 | |
| | | | 3.97 | |
| | | | 4.45 | |
| **31** | **Leucine** | δ-$CH_3$ | 0.95 | d |
| | | δ-$CH_3$ | 0.96 | d |
| | | | 1.66 | m |
| | | | 1.70 | m |
| | | | 1.73 | m |
| | | α-CH | 3.71 | |
| **32** | **Lysine** | ɣ-$CH_2$ | 1.43 | m |
| | | ɣ-$CH_2$ | 1.49 | m |
| | | δ-$CH_2$ | 1.72 | m |
| | | β-$CH_2$ | 1.88 | m |
| | | β-$CH_2$ | 1.91 | m |
| | | | 3.02 | t |
| | | | 3.74 | t |
| **33** | **Mannose** | | 4.89 | d |
| | | | 5.18 | d |
| **34** | **Methanol** | $CH_3OH$ | 3.35 | s |
| **35** | **Methionine** | | 2.15 | s |
| | | | 2.64 | t |
| **36** | **Myo-inositol** | | 3.27 | t |
| **37** | **N-acetyl-glycoprotein 1** | $NHCOCH_3$ | 2.04 | s |
| **38** | **N-acetyl-glycoprotein 2** | $NHCOCH_3$ | 2.07 | s |
| **39** | **Phenylalanine** | half β-$CH_2$ | 3.26 | |
| | | α-CH | 3.97 | |
| | | H2, H6 | 7.31 | d |
| | | H4 | 7.35 | m |
| | | H3, H5 | 7.40 | m |
| **40** | **Proline** | ɣ-$CH_2$ | 1.98 | m |
| | | ɣ-$CH_2$ | 2.01 | m |
| | | half β-$CH_2$ | 2.05 | m |
| | | half β-$CH_2$ | 2.34 | m |
| | | half δ-$CH_2$ | 3.33 | m |
| | | α-CH | 4.12 | m |
| **41** | **Succinate** | | 2.39 | s |
| **42** | **Threonine** | ɣ-$CH_3$ | 1.31 | d |
| | | α-CH | 3.55 | d |
| | | β-CH | 4.23 | m |
| **43** | **Trehalose** | | 3.40 | |
| **44** | **Tyrosine** | | 6.88 | d |
| | | H2, H6 | 7.18 | d |
| **45** | **Urea** | $NH_2C{=}ONH_2$ | 5.77 | s |
| **46** | **Valine** | CH3 | 0.98 | d |
| | | CH3 | 1.03 | d |
| | | β-CH | 2.26 | m |
| | | α-CH | 3.60 | d |
| **47** | **Xylose** | | 3.41 | |

For each metabolite, chemical group, δ in ppm relative to alanine peak at 1.475, and multiplicity of peak are presented. Singulet (s), doublet (d), doublet doublet (dd), multiplet (m).

Supplemental Table S6

**Table S6: List of compounds that are quantified in serum samples with Bruker Avance 600 MHz IVDr NMR platform, using the B.I.QUANT-PS package for metabolites and the B.I.LISA package for lipoproteins.**

| Molecule group | Compound | Class / Subclass | Abbreviation | Unit | Exclusion rationales[A] |
|---|---|---|---|---|---|
| Lipoproteins | Apolipoprotein-A1 | Total Plasma | TPA1 | mg/dL | — |
| Lipoproteins | Apolipoprotein-A1 | HDL Class | HDA1 | mg/dL | — |
| Lipoproteins | Apolipoprotein-A1 | HDL-1 Subclass | H1A1 | mg/dL | — |
| Lipoproteins | Apolipoprotein-A1 | HDL-2 Subclass | H2A1 | mg/dL | — |
| Lipoproteins | Apolipoprotein-A1 | HDL-3 Subclass | H3A1 | mg/dL | — |
| Lipoproteins | Apolipoprotein-A1 | HDL-4 Subclass | H4A1 | mg/dL | — |
| Lipoproteins | Apolipoprotein-A2 | Total Plasma | TPA2 | mg/dL | — |
| Lipoproteins | Apolipoprotein-A2 | HDL Class | HDA2 | mg/dL | — |
| Lipoproteins | Apolipoprotein-A2 | HDL-1 Subclass | H1A2 | mg/dL | — |
| Lipoproteins | Apolipoprotein-A2 | HDL-2 Subclass | H2A2 | mg/dL | — |
| Lipoproteins | Apolipoprotein-A2 | HDL-3 Subclass | H3A2 | mg/dL | — |
| Lipoproteins | Apolipoprotein-A2 | HDL-4 Subclass | H4A2 | mg/dL | — |
| Lipoproteins | Apolipoprotein-B100 | Total Plasma | TPAB | mg/dL | — |
| Lipoproteins | Apolipoprotein-B100 | VLDL Class | VLAB | mg/dL | — |
| Lipoproteins | Apolipoprotein-B100 | IDL Class | IDAB | mg/dL | — |
| Lipoproteins | Apolipoprotein-B100 | LDL Class | LDAB | mg/dL | — |
| Lipoproteins | Apolipoprotein-B100 | LDL-1 Subclass | L1AB | mg/dL | — |
| Lipoproteins | Apolipoprotein-B100 | LDL-2 Subclass | L2AB | mg/dL | — |
| Lipoproteins | Apolipoprotein-B100 | LDL-3 Subclass | L3AB | mg/dL | — |
| Lipoproteins | Apolipoprotein-B100 | LDL-4 Subclass | L4AB | mg/dL | — |
| Lipoproteins | Apolipoprotein-B100 | LDL-5 Subclass | L5AB | mg/dL | — |
| Lipoproteins | Apolipoprotein-B100 | LDL-6 Subclass | L6AB | mg/dL | — |
| Lipoproteins | Apolipoprotein-A1 / Apolipoprotein-B100 | Ratio of Apolipoproteins A1 and B100 | ABA1 | -/- | — |
| Lipoproteins | Cholesterol | Total Plasma | TPCH | mg/dL | — |
| Lipoproteins | Cholesterol | LDL | LDCH | mg/dL | — |
| Lipoproteins | Cholesterol | HDL | HDCH | mg/dL | — |
| Lipoproteins | Cholesterol | VLDL Class | VLCH | mg/dL | — |
| Lipoproteins | Cholesterol | IDL Class | IDCH | mg/dL | — |
| Lipoproteins | Cholesterol | VLDL-1 Subclass | V1CH | mg/dL | — |
| Lipoproteins | Cholesterol | VLDL-2 Subclass | V2CH | mg/dL | — |
| Lipoproteins | Cholesterol | VLDL-3 Subclass | V3CH | mg/dL | — |
| Lipoproteins | Cholesterol | VLDL-4 Subclass | V4CH | mg/dL | — |
| Lipoproteins | Cholesterol | VLDL-5 Subclass | V5CH | mg/dL | — |
| Lipoproteins | Cholesterol | LDL-1 Subclass | L1CH | mg/dL | — |
| Lipoproteins | Cholesterol | LDL-2 Subclass | L2CH | mg/dL | — |
| Lipoproteins | Cholesterol | LDL-3 Subclass | L3CH | mg/dL | — |
| Lipoproteins | Cholesterol | LDL-4 Subclass | L4CH | mg/dL | — |
| Lipoproteins | Cholesterol | LDL-5 Subclass | L5CH | mg/dL | — |
| Lipoproteins | Cholesterol | LDL-6 Subclass | L6CH | mg/dL | — |
| Lipoproteins | Cholesterol | HDL-1 Subclass | H1CH | mg/dL | — |
| Lipoproteins | Cholesterol | HDL-2 Subclass | H2CH | mg/dL | — |
| Lipoproteins | Cholesterol | HDL-3 Subclass | H3CH | mg/dL | — |
| Lipoproteins | Cholesterol | HDL-4 Subclass | H4CH | mg/dL | — |
| Lipoproteins | LDL Cholesterol / HDL Cholesterol | Ratio LDL and HDL Cholesterol | LDHD | -/- | — |
| Lipoproteins | Free Cholesterol | VLDL Class | VLFC | mg/dL | — |
| Lipoproteins | Free Cholesterol | IDL Class | IDFC | mg/dL | — |
| Lipoproteins | Free Cholesterol | LDL Class | LDFC | mg/dL | — |
| Lipoproteins | Free Cholesterol | HDL Class | HDFC | mg/dL | — |
| Lipoproteins | Free Cholesterol | VLDL-1 Subclass | V1FC | mg/dL | — |
| Lipoproteins | Free Cholesterol | VLDL-2 Subclass | V2FC | mg/dL | — |
| Lipoproteins | Free Cholesterol | VLDL-3 Subclass | V3FC | mg/dL | — |
| Lipoproteins | Free Cholesterol | VLDL-4 Subclass | V4FC | mg/dL | — |
| Lipoproteins | Free Cholesterol | VLDL-5 Subclass | V5FC | mg/dL | — |
| Lipoproteins | Free Cholesterol | LDL-1 Subclass | L1FC | mg/dL | — |
| Lipoproteins | Free Cholesterol | LDL-2 Subclass | L2FC | mg/dL | — |
| Lipoproteins | Free Cholesterol | LDL-3 Subclass | L3FC | mg/dL | — |
| Lipoproteins | Free Cholesterol | LDL-4 Subclass | L4FC | mg/dL | — |
| Lipoproteins | Free Cholesterol | LDL-5 Subclass | L5FC | mg/dL | — |
| Lipoproteins | Free Cholesterol | LDL-6 Subclass | L6FC | mg/dL | — |
| Lipoproteins | Free Cholesterol | HDL-1 Subclass | H1FC | mg/dL | — |
| Lipoproteins | Free Cholesterol | HDL-2 Subclass | H2FC | mg/dL | — |
| Lipoproteins | Free Cholesterol | HDL-3 Subclass | H3FC | mg/dL | — |
| Lipoproteins | Free Cholesterol | HDL-4 Subclass | H4FC | mg/dL | — |
| Lipoproteins | Phospholipids | VLDL Class | VLPL | mg/dL | — |
| Lipoproteins | Phospholipids | IDL Class | IDPL | mg/dL | — |
| Lipoproteins | Phospholipids | LDL Class | LDPL | mg/dL | — |
| Lipoproteins | Phospholipids | HDL Class | HDPL | mg/dL | — |
| Lipoproteins | Phospholipids | VLDL-1 Subclass | V1PL | mg/dL | — |
| Lipoproteins | Phospholipids | VLDL-2 Subclass | V2PL | mg/dL | — |
| Lipoproteins | Phospholipids | VLDL-3 Subclass | V3PL | mg/dL | — |
| Lipoproteins | Phospholipids | VLDL-4 Subclass | V4PL | mg/dL | — |
| Lipoproteins | Phospholipids | VLDL-5 Subclass | V5PL | mg/dL | — |
| Lipoproteins | Phospholipids | LDL-1 Subclass | L1PL | mg/dL | — |
| Lipoproteins | Phospholipids | LDL-2 Subclass | L2PL | mg/dL | — |
| Lipoproteins | Phospholipids | LDL-3 Subclass | L3PL | mg/dL | — |
| Lipoproteins | Phospholipids | LDL-4 Subclass | L4PL | mg/dL | — |

| | | | | | |
|---|---|---|---|---|---|
| Lipoproteins | Phospholipids | LDL-5 Subclass | L5PL | mg/dL | — |
| Lipoproteins | Phospholipids | LDL-6 Subclass | L6PL | mg/dL | — |
| Lipoproteins | Phospholipids | HDL-1 Subclass | H1PL | mg/dL | — |
| Lipoproteins | Phospholipids | HDL-2 Subclass | H2PL | mg/dL | — |
| Lipoproteins | Phospholipids | HDL-3 Subclass | H3PL | mg/dL | — |
| Lipoproteins | Phospholipids | HDL-4 Subclass | H4PL | mg/dL | — |
| Lipoproteins | Triglycerides | Total Plasma | TPTG | mg/dL | — |
| Lipoproteins | Triglycerides | VLDL Class | VLTG | mg/dL | — |
| Lipoproteins | Triglycerides | IDL Class | IDTG | mg/dL | — |
| Lipoproteins | Triglycerides | LDL Class | LDTG | mg/dL | — |
| Lipoproteins | Triglycerides | HDL Class | HDTG | mg/dL | — |
| Lipoproteins | Triglycerides | VLDL-1 Subclass | V1TG | mg/dL | — |
| Lipoproteins | Triglycerides | VLDL-2 Subclass | V2TG | mg/dL | — |
| Lipoproteins | Triglycerides | VLDL-3 Subclass | V3TG | mg/dL | — |
| Lipoproteins | Triglycerides | VLDL-4 Subclass | V4TG | mg/dL | — |
| Lipoproteins | Triglycerides | VLDL-5 Subclass | V5TG | mg/dL | — |
| Lipoproteins | Triglycerides | LDL-1 Subclass | L1TG | mg/dL | — |
| Lipoproteins | Triglycerides | LDL-2 Subclass | L2TG | mg/dL | — |
| Lipoproteins | Triglycerides | LDL-3 Subclass | L3TG | mg/dL | — |
| Lipoproteins | Triglycerides | LDL-4 Subclass | L4TG | mg/dL | — |
| Lipoproteins | Triglycerides | LDL-5 Subclass | L5TG | mg/dL | — |
| Lipoproteins | Triglycerides | LDL-6 Subclass | L6TG | mg/dL | — |
| Lipoproteins | Triglycerides | HDL-1 Subclass | H1TG | mg/dL | — |
| Lipoproteins | Triglycerides | HDL-2 Subclass | H2TG | mg/dL | — |
| Lipoproteins | Triglycerides | HDL-3 Subclass | H3TG | mg/dL | — |
| Lipoproteins | Triglycerides | HDL-4 Subclass | H4TG | mg/dL | — |
| Lipoproteins | Particle Number | LDL-1 | L1PN | nmol/L | — |
| Lipoproteins | Particle Number | Apolipoprotein-B100 carrying particles | TBPN | nmol/L | — |
| Lipoproteins | Particle Number | VLDL | VLPN | nmol/L | — |
| Lipoproteins | Particle Number | IDL | IDPN | nmol/L | — |
| Lipoproteins | Particle Number | LDL | LDPN | nmol/L | — |
| Lipoproteins | Particle Number | LDL-2 | L2PN | nmol/L | — |
| Lipoproteins | Particle Number | LDL-3 | L3PN | nmol/L | — |
| Lipoproteins | Particle Number | LDL-4 | L4PN | nmol/L | — |
| Lipoproteins | Particle Number | LDL-5 | L5PN | nmol/L | — |
| Lipoproteins | Particle Number | LDL-6 | L6PN | nmol/L | — |
| Metabolites | 2-Aminobutyric acid | — | — | mmol/L | Missing values: 74%/ median ρ= 0% |
| Metabolites | 2-Hydroxybutyric acid | — | — | mmol/L | Missing values: 75%/ median ρ= 0% |
| Metabolites | 2-Oxoglutaric acid | — | α-KG | mmol/L | Missing values: 65%/ median ρ= 0% |
| Metabolites | 3-Hydroxybutyric acid | — | BHB | mmol/L | — |
| Metabolites | Acetic acid | — | — | mmol/L | — |
| Metabolites | Acetoacetic acid | — | AcAc | mmol/L | — |
| Metabolites | Acetone | — | — | mmol/L | — |
| Metabolites | Alanine | — | Ala | mmol/L | — |
| Metabolites | Asparagine | — | Asn | mmol/L | Missing values: 100%/ median ρ= 0% |
| Metabolites | Betaine | — | — | mmol/L | — |
| Metabolites | Ca-EDTA | — | — | mmol/L | Median ρ= 0% |
| Metabolites | Choline | — | — | mmol/L | Missing values: 54%/ median ρ= 0% |
| Metabolites | Citric acid | — | — | mmol/L | — |
| Metabolites | Creatine | — | — | mmol/L | — |
| Metabolites | Creatinine | — | — | mmol/L | — |
| Metabolites | D-Galactose | — | — | mmol/L | Missing values: 100%/ median ρ= 0% |
| Metabolites | Dimethylsulfone | — | DMSO2 | mmol/L | — |
| Metabolites | Ethanol | — | EtOH | mmol/L | Potential contaminant |
| Metabolites | Formic acid | — | — | mmol/L | — |
| Metabolites | Glucose | — | Glc | mmol/L | — |
| Metabolites | Glutamic acid | — | Glu | mmol/L | Missing values: 38%/ median ρ= 21% |
| Metabolites | Glutamine | — | Gln | mmol/L | — |
| Metabolites | Glycerol | — | Glyc | mmol/L | Missing values: 52%/ median ρ= 0% |
| Metabolites | Glycine | — | Gly | mmol/L | — |
| Metabolites | Histidine | — | His | mmol/L | — |
| Metabolites | Isoleucine | — | Ile | mmol/L | — |
| Metabolites | K-EDTA | — | — | mmol/L | Quality control variables |
| Metabolites | Lactic acid | — | Lac | mmol/L | — |
| Metabolites | Leucine | — | Leu | mmol/L | — |
| Metabolites | Lysine | — | Lys | mmol/L | Median ρ= 64% |
| Metabolites | Methionine | — | Met | mmol/L | — |
| Metabolites | N,N-Dimethylglycine | — | DMG | mmol/L | — |
| Metabolites | Ornithine | — | — | mmol/L | Missing values: 41%/ median ρ= 11% |
| Metabolites | Phenylalanine | — | Phe | mmol/L | — |
| Metabolites | Proline | — | Pro | mmol/L | Missing values: 81%/ median ρ= 0% |
| Metabolites | Pyruvic acid | — | Pyr | mmol/L | — |
| Metabolites | Sarcosine | — | — | mmol/L | Median ρ= 74% |
| Metabolites | Succinic acid | — | Succ | mmol/L | — |
| Metabolites | Threonine | — | Thr | mmol/L | Missing values: 42%/ |

| | | | | | |
|---|---|---|---|---|---|
| | | | | | median ρ= 63% |
| Metabolites | Trimethylamine-N-oxide | — | TMAO | mmol/L | — |
| Metabolites | Tyrosine | — | Tyr | mmol/L | — |
| Metabolites | Valine | — | Val | mmol/L | — |

[A] Several metabolites were excluded from statistical analysis based on proportion of missing values >30%, median sigCorr ρ < 75%, quality control variables or potential contaminant. LDL, low-density lipoprotein; HDL, high-density lipoprotein; VLDL, very low-density lipoprotein; and IDL, intermediate-density lipoprotein.

### Supplemental figures

Supplemental Figure S1

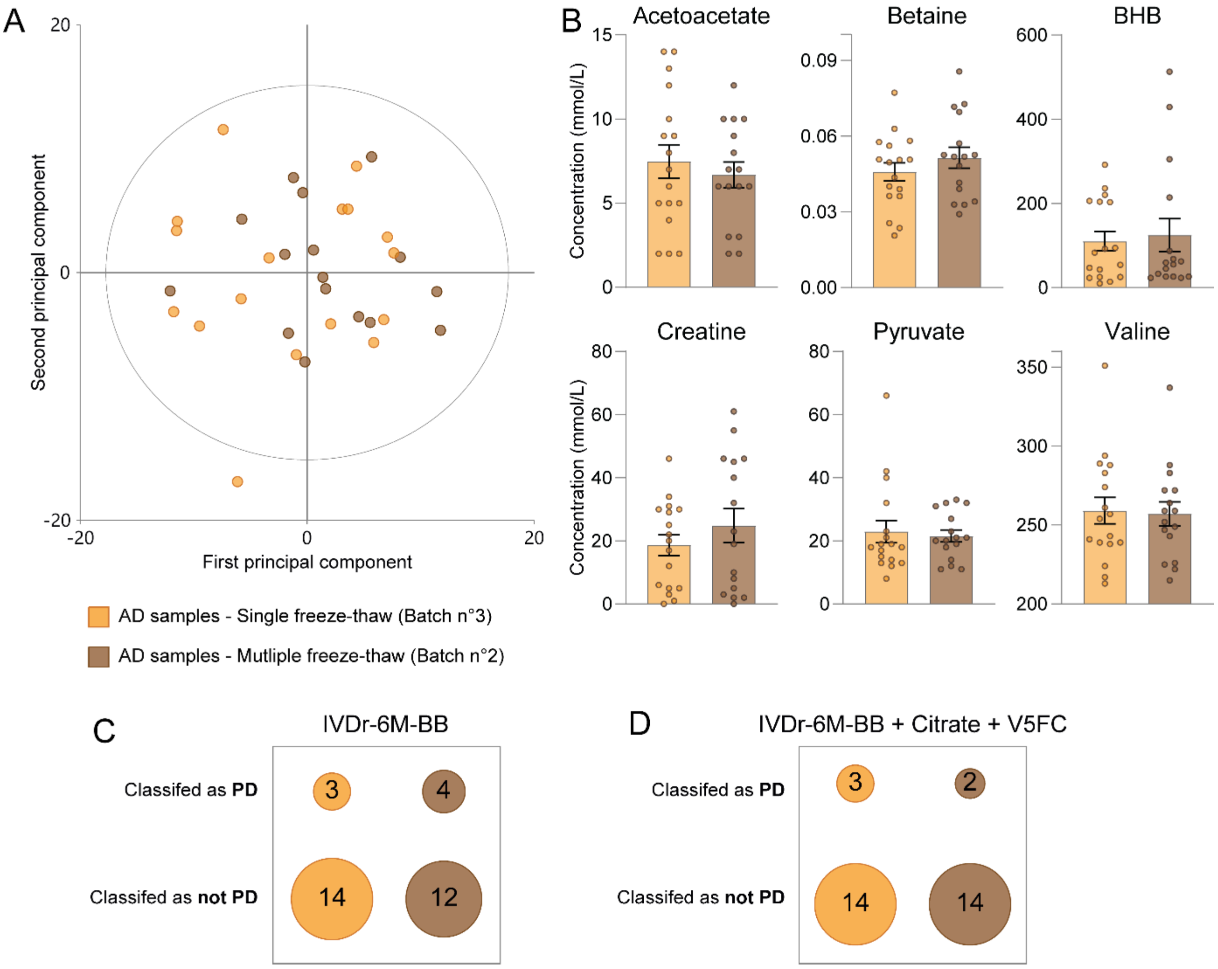

**<u>Figure S1</u>: AD metabolic profile and classification remain unaffected by multiple freeze–thaw cycles.** (**A**) Score of the Principal Component Analysis (PCA) model plotted versus the 2 first principal components. The PCA was built with absolute concentrations of metabolites and lipoproteins in the batches of AD patients: batch n°3 with a single freeze-thaw (n=17, orange) and batch n°2 with multiple freeze-thaw (n=16, brown). R2X[1]=0.334; R2X[2]=0.242; 4 principal components. There is no separation between the two groups. (**B**) Similar concentration (mmol/L) of the six core metabolites in the batches 2 and 3 of AD samples (n=16 and n=17; respectively). Mean ± SEM. All p-values are non-significant (t-test). (**C**) Classification table of AD cases of the batch 2 and 3 based on their IVDr-6M-BB scores (threshold 0.5100) and (D) on their IVDr-6M-BB+citrate+V5FC scores (threshold 0.4937).

Supplemental Figure S2

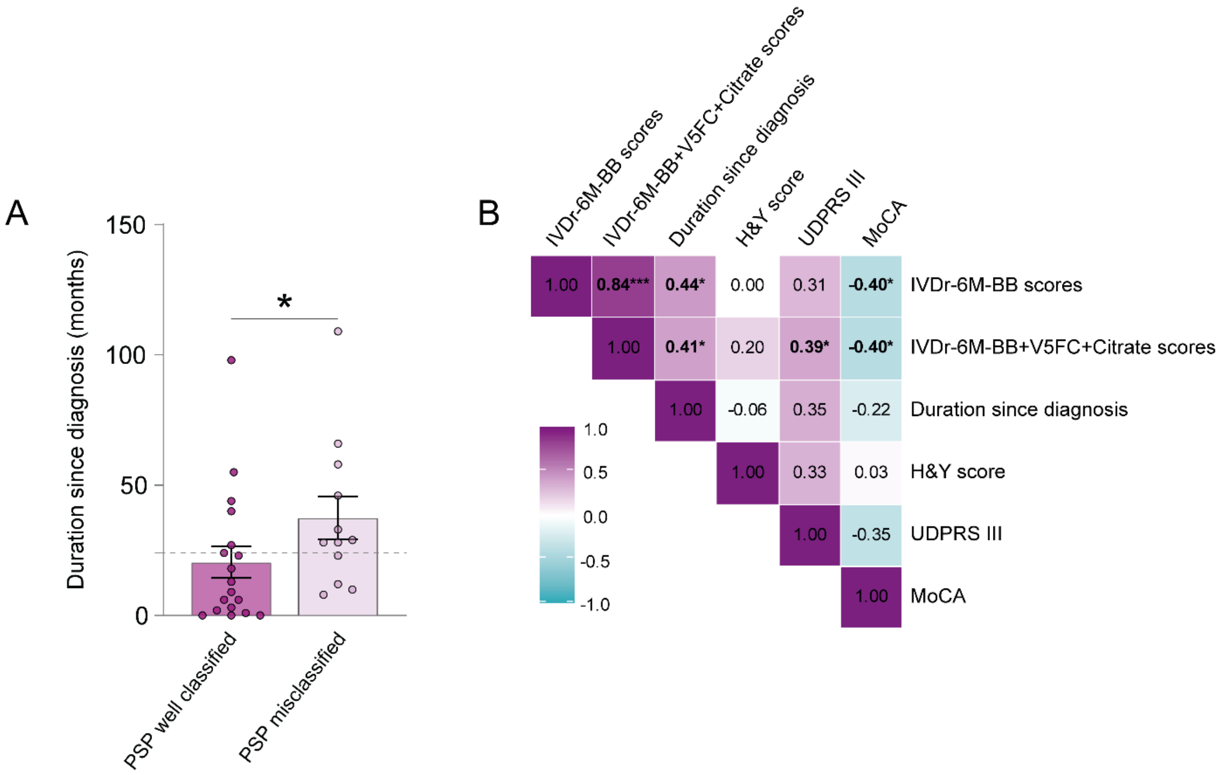

**Figure S2: Misclassified PSP patients with the IVDr biomarkers are associated with more advanced pathology.** (**A**) Duration of pathology since diagnosis (months) in PSP patients correctly classified (n=18) and misclassified (n=12) using the IVDr-6M-BB. Dotted line: 2 years (24 months) after diagnosis. Mean ± SEM. Mann-Whitney tests (*$p \leq 0.05$). (**B**) Correlation matrix of the different biomarker and disease characterization scores in the PSP dataset. Magnitude of each correlation is displayed in terms of colour, with Spearman's ρ and the corresponding p-value indicated for each pairwise comparison. (*$p \leq 0.05$; **$p \leq 0.01$; ***$p \leq 0.001$). *UPDRS: Unified Parkinson's Disease Rating Scale. H&Y score: Hoehn & Yahr score. MoCA: Montreal Cognitive Assessment*

Supplemental Figure S3

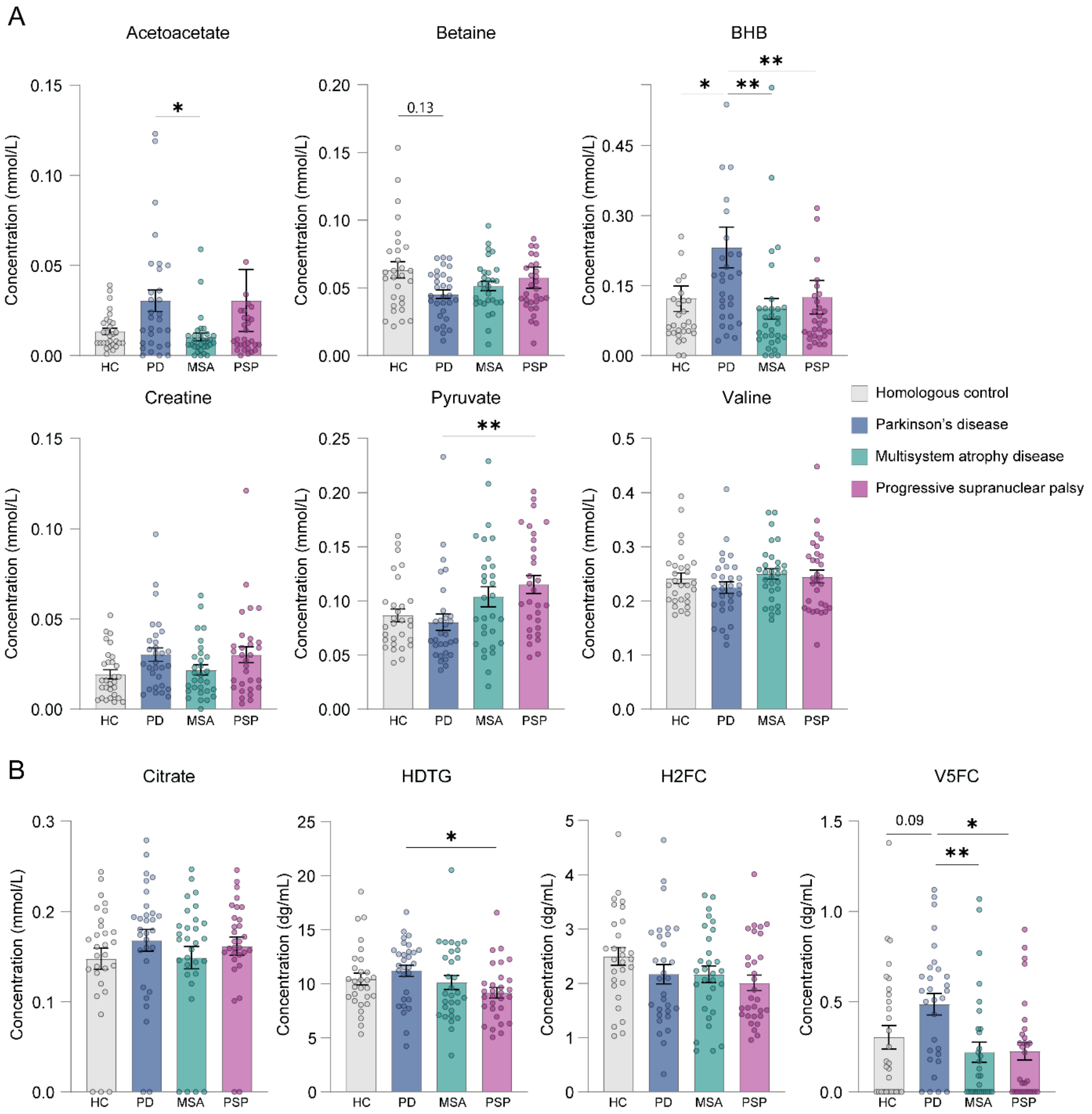

**Figure S3: Metabolic alterations between HC, PD, MSA, and PSP cases in the core metabolites and the top 4 candidate metabolites to improve IVDr-6M-BB performance. (A and B)** Quantification of (**A**) the six core metabolites and (**B**) the top 4 candidate metabolites (citrate, HDTG, H2FC and V5FC) in HC (n=29, grey), PD (n=30, dark blue), MSA (n=30, turquoise), and PSP (n=30, purple). All data are represented as mean ± SEM, Kruskal-Wallis tests followed by Dunn's post-hoc tests for multiple comparisons. (*$p \leq 0.05$; **$p \leq 0.01$; ***$p \leq 0.001$; ****$p \leq 0.0001$).

Supplemental Figure S4

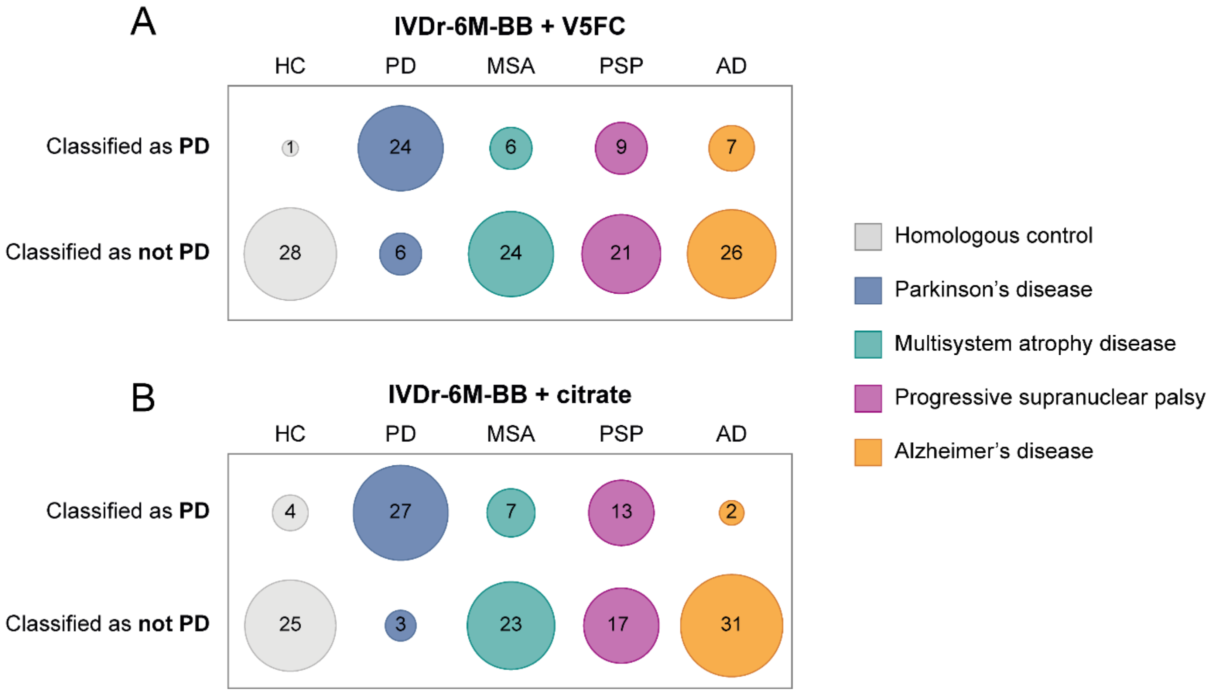

**<u>Figure S4</u>: Accuracies associated with the IVDr-6M-BB added with V5FC or citrate. (A)** Classification table of HC (n=29), PD (n=30), MSA (n=30), PSP (n=30), and AD (n=33) cases based on the regression model combining the 6 core metabolites plus V5FC. The resulting scores were used to classify the samples as "PD" or "no PD" with the threshold of 0.646. Accuracies: HC, 96.6%; PD, 80.0%; MSA, 80.0%; PSP, 70.0%; and AD, 78.8%. Overall accuracy: 80.9% (81.5 % considering only parkinsonian syndromes and HC). (**B**) Classification table of HC (n=29), PD (n=30), MSA (n=30), PSP (n=30), and AD (n=33) cases based on the regression model combining the 6 core metabolites plus citrate. The resulting scores were used to classify the samples as "PD" or "no PD" with the threshold of 0.471. Accuracies: HC, 86.2%; PD, 90.0%; MSA, 76.7%; PSP, 56.7%; and AD, 93.9%. Overall accuracy: 80.9% (77.3% considering only parkinsonian syndromes and HC).

Supplemental Figure S5

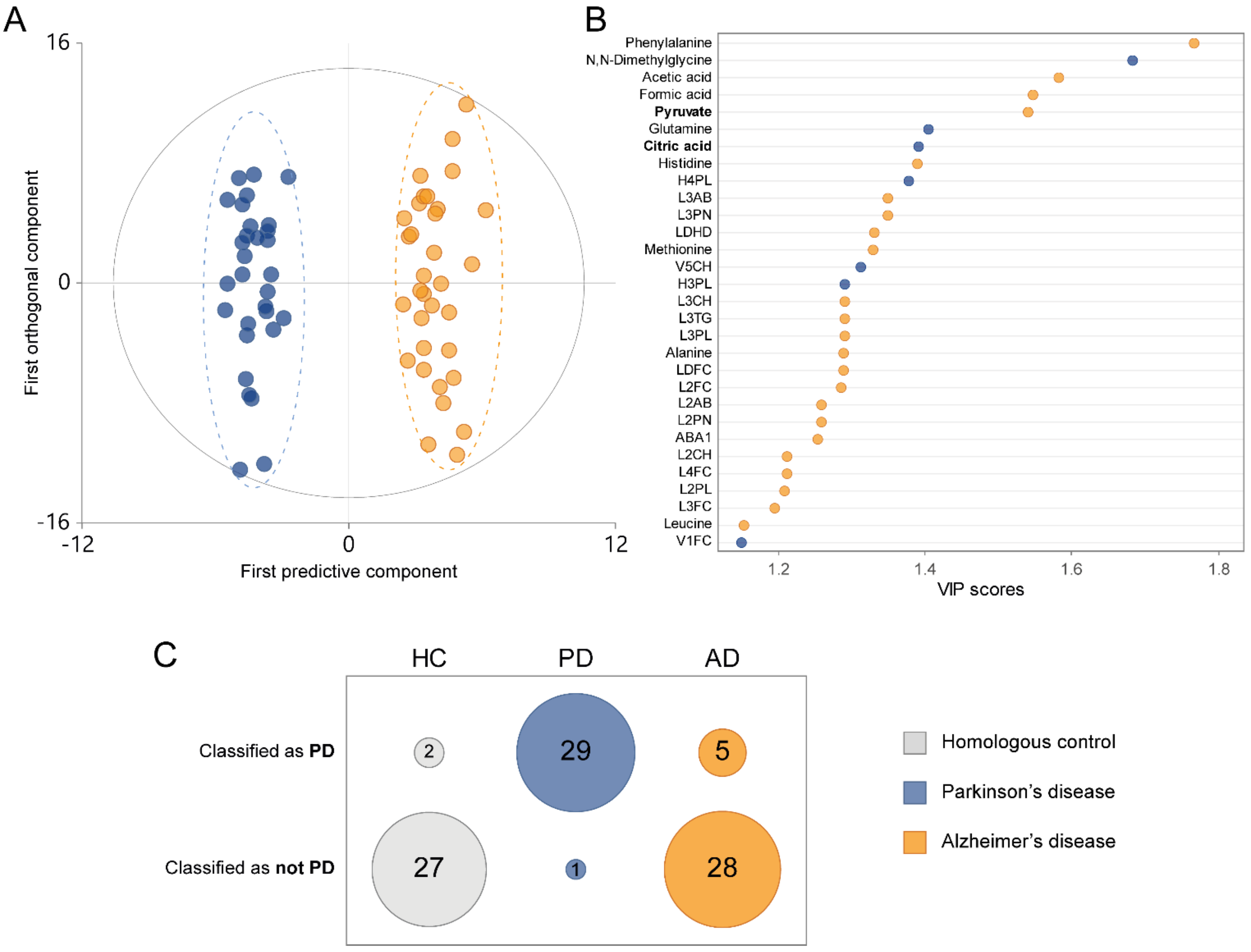

**Figure S5: Citrate is part of the top-10 VIP for discriminating PD from AD, and its added value in the IVDr-6M-BB, along with V5FC, helps to maintain high level of performance for the classification of AD cases**. (**A** and **B**) OPLS-DA model built with the absolute concentrations of metabolites and lipoproteins in PD (n=30, dark blue) and AD (n=33, orange). R2Y=0.961; Q2=0.931; 1 predictive and 4 orthogonal components; CV-ANOVA, $p = 1.24\times10^{-26}$ (**A**) Score plot of the individuals based on the first predictive and the first orthogonal components. (**B**) Top-30 VIP plot, with the pyruvate and citrate in bold. Dark blue/orange: metabolites significantly increased in PD/AD samples (Mann-Whitney tests, detailed *p* values in Supplementary data, Table S5). (**C**) Classification table of HC (n=29), PD (n=30), and AD (n=33) cases based on the regression model combining the 6 core metabolites plus citrate and V5FC. The resulting scores were used to classify the samples as "PD" or "no PD" with the threshold of 0.4937. Accuracies: PD, 96.7%; HC, 93.1%; and AD, 84.8%.

Supplemental Figure S6

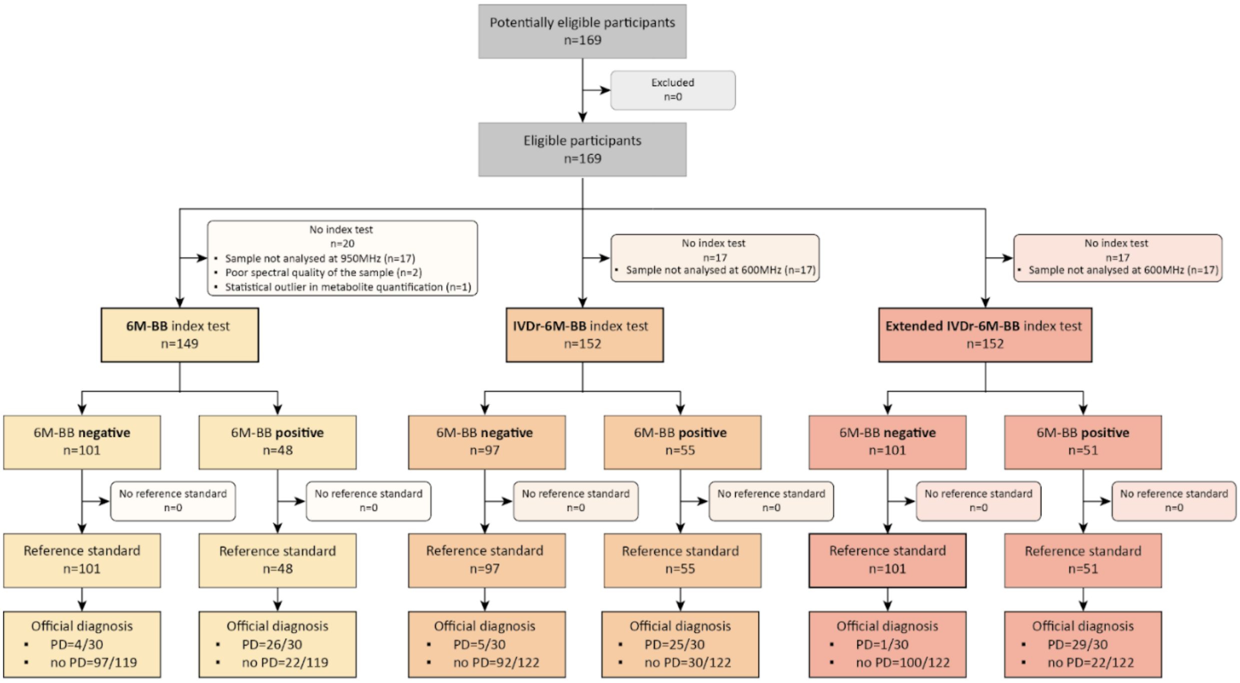

**Figure S6: Flow chart of this study according to STARD 2015 guidelines**. STARD: Standards for Reporting of Diagnostic Accuracy Studies
