## Supplementary figures and images for "Toward clinical implementation of a metabolic blood biomarker for Parkinson’s disease differential diagnosis"

### Graphical Abstract

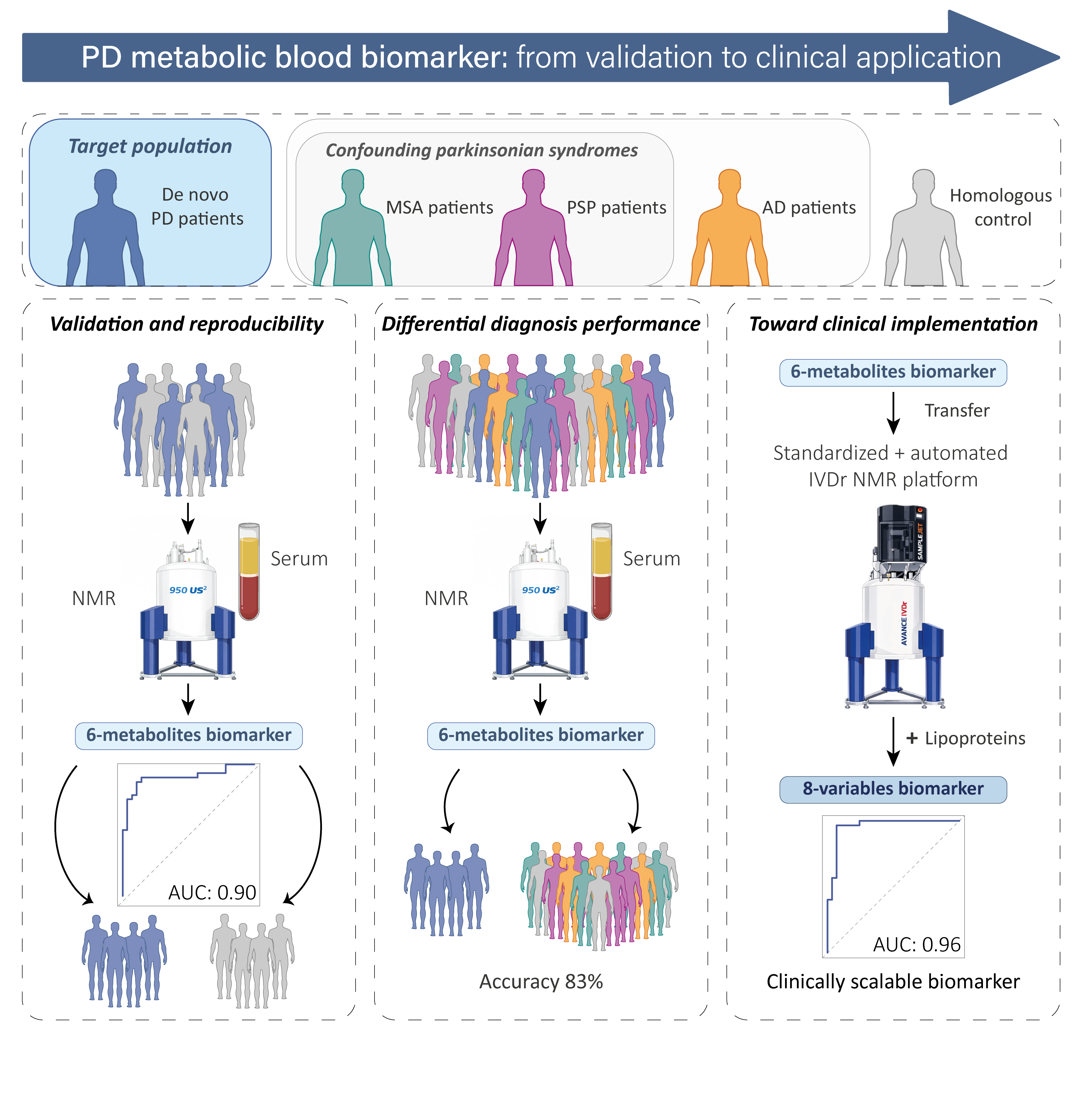
